# Robust uncertainty quantification in popular estimators of the instantaneous reproduction number

**DOI:** 10.1101/2024.10.22.24315918

**Authors:** Nicholas Steyn, Kris V Parag

**Affiliations:** Department of Statistics, University of Oxford, Oxford, United Kingdom; MRC Centre for Global Infectious Disease Analysis, Imperial College London, London, United Kingdom

## Abstract

The instantaneous reproduction number (*R*_*t*_) is a key measure of the rate of spread of an infectious disease. Correctly quantifying uncertainty in *R*_*t*_ estimates is crucial for making well-informed decisions. Popular *R*_*t*_ estimators leverage smoothing techniques to distinguish signal from noise. Examples include EpiEstim and EpiFilter, which are both controlled by a “smoothing parameter” that is traditionally selected by users. We demonstrate that the values of these smoothing parameters are unknown, vary markedly with epidemic dynamics, and show that data-driven smoothing is crucial for accurate uncertainty quantification of *R*_*t*_ estimates. We derive model likelihoods for the smoothing parameters in both EpiEstim and EpiFilter and develop a Bayesian framework to automatically marginalise these parameters when fitting to epidemiological time-series data. This yields novel marginal posterior predictive distributions which prove integral to rigorous model evaluation. Applying our methods, we find that default parameterisations of these widely-used estimators can negatively impact *R*_*t*_ inference, delaying detection of epidemic growth, and misrepresenting uncertainty (typically producing overconfident estimates), with implications for public health decision-making. Our extensions mitigate these issues, provide a principled approach to uncertainty quantification, improve the robustness of real-time *R*_*t*_ inference, and facilitate model comparison using observable quantities.

---

The instantaneous reproduction number *R*_*t*_, defined as the average number of infections generated per primary infection at time *t*, is a popular measure of epidemic spread [1]. A value of *R*_*t*_ *<* 1 indicates a declining epidemic while *R*_*t*_ *>* 1 indicates a growing epidemic. This quantity is useful for policymakers as it gives the change in transmission required to control the epidemic, informing decisions about public health interventions [2, 3, 4, 5, 6]. As a stark example, in June 2020, an *R*_*t*_ estimate of 1.01 was used to justify continued school closures in Greater Manchester, England [7, 8]. Additionally, estimates of *R*_*t*_ are also used for forecasting, scenario analysis, and understanding the impact of interventions [9].

Many models exist to estimate *R*_*t*_, including compartmental models, agent-based models, and the renewal-model [10], the latter of which underlies most contemporary methods for real-time estimation of *R*_*t*_ from reported case time-series, including EpiEstim [11] and EpiFilter [12]. Reflecting a community drive to improve existing methods and make public health pipelines more reliable, we focus on quantifying uncertainty in these two models, although our approach generalises to any real-time model.

Correct quantification of the uncertainty of *R*_*t*_ is crucial for making well-informed decisions. It is expected that the true value of *R*_*t*_ should fall within a 95% credible interval 95% of the time. Undercoverage occurs when this happens less often than expected, leading to over-confident and biased decision-making. Overcoverage occurs when this happens more often than expected, leading to under-confident and highly uncertain decision-making. A model that produces correct coverage is termed well-calibrated.

Despite the importance of uncertainty quantification, calibration is often neglected in epidemiological models. For example, during the COVID-19 pandemic, SPI-M in England pooled estimates from multiple groups to produce consensus *R*_*t*_ estimates. However, pooling proved difficult due to models “*providing estimates with lower levels of uncertainty that are not fully accounting for inherent uncertainties*” [13]. Even when correct coverage is explicitly targeted, epidemic models frequently fail to achieve it. For instance, nearly all models submitted to the *open challenge to advance probabilistic forecasting for dengue epidemics* [14] produced over-confident predictions across various forecasting targets. A baseline model (included for comparison) demonstrated superior calibration compared to all 16 submitted models when predicting the peak of the epidemic.

Smoothing assumptions, which include penalized likelihoods [15], piecewise constant/trailing window models [11, 16, 17, 18], and latent-space models [12, 19, 20, 21], are a key source of model miscalibration [22]. Oversmoothed estimates result in delayed and overconfident estimates, while undersmoothed estimates are noisy and lack precision. Even with perfect case reporting (i.e. no observation noise), inherent stochasticity in the transmission of infectious diseases necessitates the use of smoothing to distinguish signal from noise. All popular estimators of *R*_*t*_ employ some type of smoothing.

Despite the importance and ubiquity of these assumptions, the philosophical and practical treatment of smoothing parameters varies by method. Some methods treat these parameters as unknown quantities to be estimated alongside *R*_*t*_ [19, 22, 23, 24], while others treat the choice of smoothing parameter(s) as a model selection problem and seek to find a fixed, optimal parameter value [22, 25, 26]. Some methods allow the user to choose their own values or provide heuristic default values [11, 15, 24].

We argue that, because the true dynamics of *R*_*t*_ are always unknown and depend on both the pathogen and the population in which the pathogen is spreading, uncertainty about the nature of these dynamics should not be ignored. This uncertainty factors in both the choice of the dynamic model itself (structural uncertainty), and the parameters associated with the chosen dynamic model (parametric uncertainty). We focus on parametric uncertainty in this paper, which on its own can cause substantial model miscalibration, and demonstrate that correct marginalisation of smoothing parameters generally improves model robustness. This result holds even if the structure of the dynamic model does not accurately reflect reality, as the increased flexibility of the model allows it to better adapt to the data.

In this paper, we derive novel likelihoods for smoothing parameters in two popular models: Cori et al. [11] and Parag [12], commonly known as EpiEstim and EpiFilter respectively^1^. We use these likelihoods to marginalise the smoothing parameters, presenting estimates of *R*_*t*_ that appropriately account for uncertainty in these parameters. We also derive predictive posterior distributions and demonstrate their use in model comparison via the continuous ranked probability score (CRPS) [27], emphasising the importance of model comparison using observable quantities. We validate our methods on both simulated data, where model estimates can be compared to ground truths, and real-world data, where we investigate the practical implications on decision-making during the COVID-19 pandemic in New Zealand. For each dataset considered, we fit four models: EpiEstim and EpiFilter with default parameters and with our marginalised algorithm.

## Methods

We summarise our methods here but provide full mathematical derivations and technical details in supplementary section 1.

### Background

Both EpiEstim and EpiFilter leverage the *Poisson renewal model* for *R*_*t*_ estimation. Letting *C*_*s*_ be the number of cases reported at time *s* and *w*_*u*_ be the probability that a secondary case is reported *u* days after the primary case (often approximated by the serial interval). The *total infectiousness* is defined as 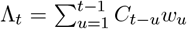 Given Λ and the current value of *R*_*t*_, the Poisson renewal model considers the number of cases at time *t* to be Poisson distributed:

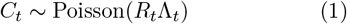

EpiEstim assumes that, on each day *t, R*_*t*_ has been fixed for a trailing window of *k* days. Larger values of *k* imply that *R*_*t*_ has been fixed for a longer period, resulting in smoother estimates. The likelihood of observing cases between time-steps *t* − *k* + 1 and *t* (denoted *C*_*t*−*k*+1:*t*_) is the product of the daily Poisson likelihoods (equation 1) along the trailing window. A conjugate Gamma(*α, β*) prior distribution is assumed for *R*_*t*_, resulting in a Gamma(*α*_*t,k*_, *β*_*t,k*_) posterior distribution for *R*_*t*_ given *C*_1:*t*_, where the shape-parameter *α*_*t,k*_ and rate-parameter *β*_*t,k*_ are:

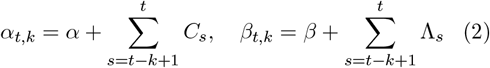

Alternatively, EpiFilter assumes that *R*_*t*_ follows a Gaussian random walk with standard deviation equal to 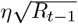 Larger values of *η* allow *R*_*t*_ to vary faster, resulting in less smooth estimates. A grid-approximation to the exact Bayesian filtering equations is used to derive the posterior distribution of *R*_*t*_ given *C*_1:*t*_. While EpiFilter also allows for the estimation of the smoothing distribution (*R*_*t*_ given past and future data), we focus on real-time estimation (*R*_*t*_ given data up to time *t*). Further discussion is included in supplementary section 2.

### Model likelihoods

We use the same framework to derive likelihoods for the smoothing parameters of both methods. Model-specific derivations are included in supplementary section 1. Letting *θ* denote an arbitrary smoothing parameter, we begin with the predictive decomposition of the likelihood:

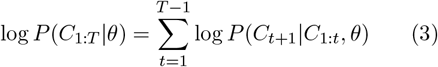

The one-step-ahead-likelihood can be written as:

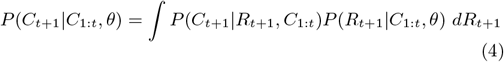

where *P* (*C*_*t*+1_|*R*_*t*+1_, *C*_1:*t*_) is the renewal model (equation 1). Thus, deriving model likelihoods relies on the derivation of the one-step-ahead predictive distribution for *R*_*t*+1_: *P* (*R*_*t*+1_|*C*_1:*t*_, *θ*).

For EpiEstim, *R*_*t*_ depends on reported cases only on days *t*−*k* +1 to *t*, however the predictive distribution explicitly ignores data on day *t*, so EpiEstim’s predictive distribution for *R*_*t*_ is Gamma-distributed with shape *α*_*t*−1,*k*−1_ and rate *β*_*t*−1,*k*−1_. In this case, equation 4 is a Gamma-Poisson mixture, hence *C*_*t*+1_|*C*_1:*t*_ follows a negative binomial distribution with parameters *r* = *α* and 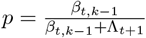 The likelihood of EpiEstim’s *k* is then the sum of log-negative binomial probability mass functions for each day *t*.

For EpiFilter, the predictive distribution of *R*_*t*+1_ is a by-product of the Bayesian filtering equations, found by propagating the estimated distribution of *R*_*t*_ given *C*_1:*t*_ forward according to the assumed Gaussian random walk. The one-step-ahead likelihood of *C*_*t*+1_ is found by taking a weighted average of the Poisson likelihood of *C*_*t*+1_ with respect to the predictive distribution of *R*_*t*+1_|*C*_1:*t*_.

Figure 1 provides a schematic of the model likelihood calculation. Implementations of these methods, alongside worked examples, are provided in the GitHub repository.

**Figure 1:**
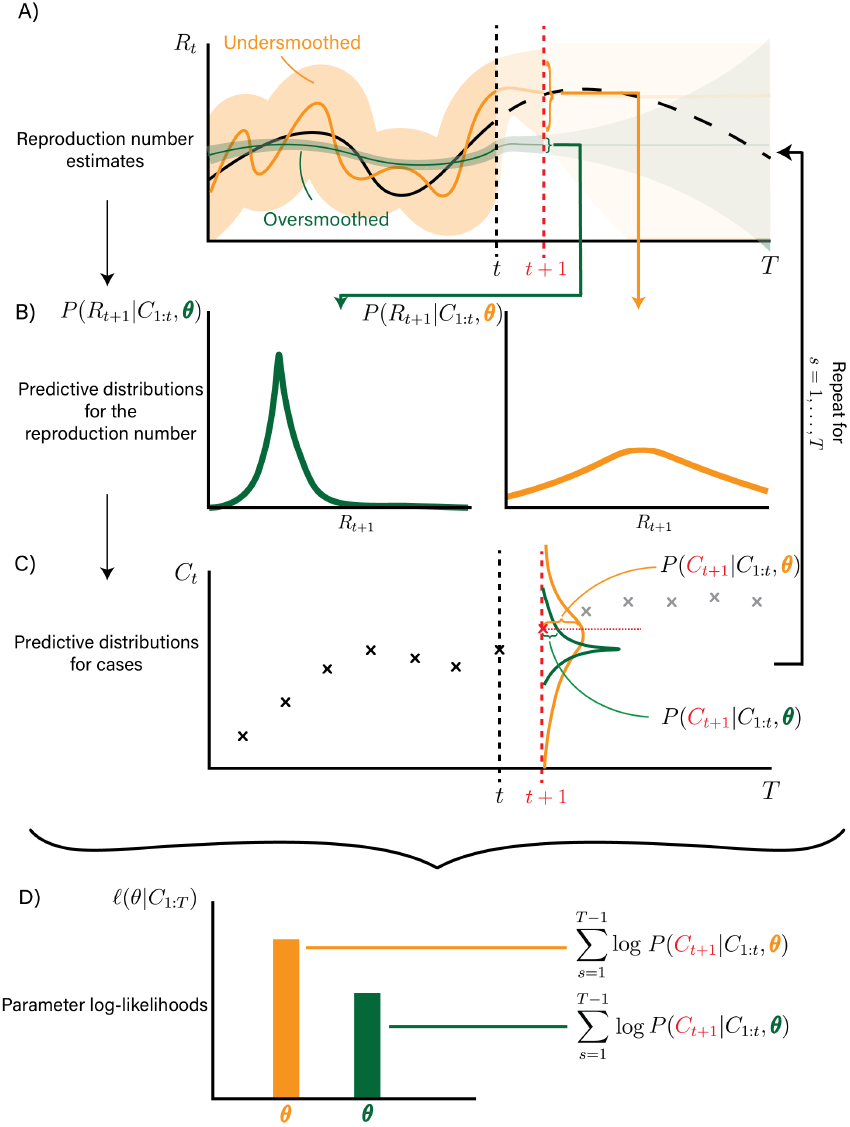
Schematic demonstrating the derivation of parameter likelihoods using the predictive decomposition (equation 3). *R*_*t*_ estimates are projected forward one time-step according to the relevant dynamic model (panel A), giving predictive distributions for *R*_*t*+1_ (panel B). These are combined with the renewal model to give the probability of observing *C*_*t*+1_ conditional on past data *C*_1:*t*_ and the chosen smoothing parameter *θ* (panel C). In this timestep, the undersmoothed model is more likely than the oversmoothed model. Repeating this process and summing the log-predictive probabilities for all timesteps produces the model likelihood (Panel D). These log-likelihoods are later used to marginalise out uncertainty about *θ*, allowing for robust reporting of uncertainty about *R*_*t*_. The form of the *R*_*t*_ estimates and predictive distributions for *R*_*t*_ depend on the specific model.

### Posterior distributions

These methods admit log-likelihoods for *k* and *η* given *C*_1:*t*_, denoted 𝓁(*k*|*C*_1:*t*_) and 𝓁(*η*|*C*_1:*t*_). We use these to derive posterior distributions (denoted *P* (*k*|*C*_1:*t*_) and *P* (*η*|*C*_1:*t*_)), typically using a uniform prior distribution over *k* ∈ {1, 2, …, 30} (covering daily to monthly dependence) and *η* ∈ [0, 1] (covering no noise to Poisson-type diffusion).

We are ultimately interested in estimates of *R*_*t*_ that account for uncertainty about *k* or *η*. To achieve this, we marginalise these parameters from the posterior distribution of *R*_*t*_, a procedure that is rare in the literature. For EpiEstim, we leverage the discrete nature of *k* to write exactly:

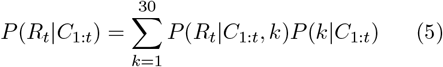

whereas for EpiFilter, we use a grid-approximation (*η* ∈ E, supplementary section 1.1.2):

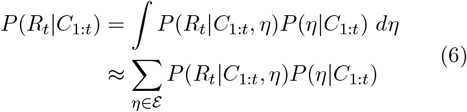

We can also marginalise the smoothing parameter from the predictive distributions for *C*_*t*_ (equation 4). This generates marginal one-step-ahead predictive distributions for *C*_*t*_ under both models, useful for model comparison and probabilistic forecasting.

### Model evaluation

We argue that a “good” model is one that maximises precision (i.e. minimises the width of uncertainty intervals), subject to being well-calibrated [27]. Choosing the “best” model thus involves a trade-off: how much miscoverage are we willing to accept in exchange for more precise estimates? We use the Continuous Ranked Probability Score (CRPS), which measures the distance between the estimated predictive distribution and the empirical distribution of the data, to quantify this trade-off. Smaller distances signify closer alignment between the model’s predictive uncertainty and observed data variability. Full details of the CRPS calculation are provided in supplementary section 1.4.

### Data

We test our methods on three simulated datasets, each assuming a different dynamic model for *R*_*t*_: a Gaussian random walk (matching the dynamic model assumed by EpiFilter), a sinusoidal curve, and a step-change model. These models cover a range of smooth to sharp changes in *R*_*t*_. We also compare model outputs on real-world data from the COVID-19 pandemic in New Zealand [28], chosen as an example of high-quality data with limited reporting biases. We explicitly relate real-world decision-making to the inferences made by our models.

A common serial interval from the COVID-19 literature, a Gamma distribution with a mean of 6.5 days and standard deviation of 4.2 days [29, 30], is used for the simulation study, while a Weibull distribution with a mean of 5.0 days and standard deviation of 1.9 days is used when fitting to real-world data, matching the serial interval used in official modelling [31].

Further information on simulated data is provided in supplementary section 3. We also develop additional marginalisation routines to handle uncertainty in serial intervals, and test sensitivity to biases in the serial interval, in supplementary section 4.

## Results

### Simulation study

Fitting EpiEstim and EpiFilter to a single realisation from each of the three simulated epidemics (figure 2) demonstrates that default parameterisations of both models result in oversmoothed estimates of *R*_*t*_, relative to the level of smoothing estimated from the data. The exception is EpiFilter in the random walk simulation, where the true value of *η* is deliberately chosen to match EpiFilter’s default *η* = 0.1.

**Figure 2:**
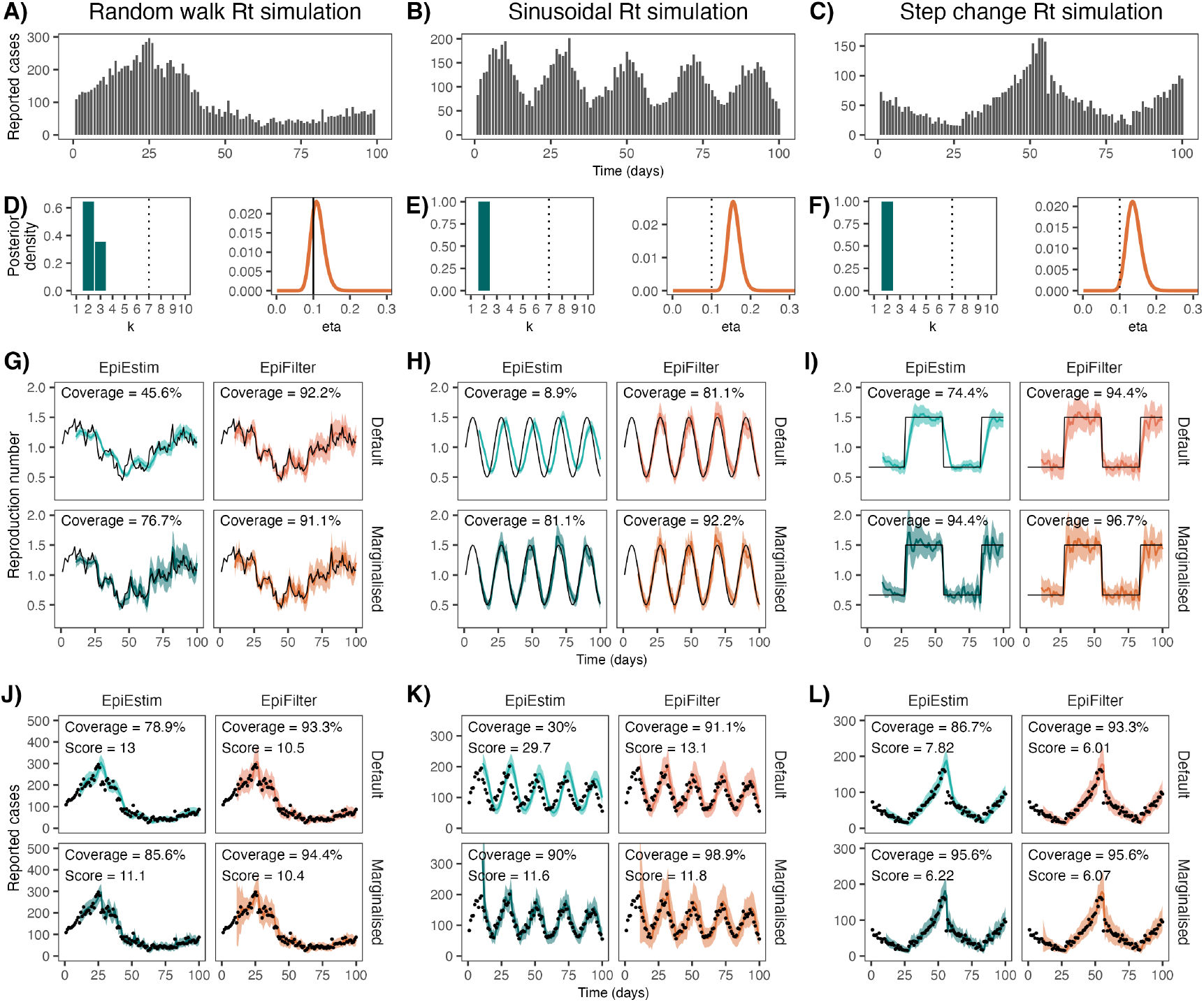
Simulated case data (A, B, C), posterior distributions for smoothing parameters at *t* = 100 (D, E, F), estimates of *R*_*t*_ (G, H, I), and estimates of predictive cases (J, K, L) for one realisation of each simulated epidemic. Estimates of *R*_*t*_ and predictive cases are shown for all four models (default EpiEstim, default EpiFilter, marginalised EpiEstim, marginalised EpiFilter). The first column (A, D, G, J) shows results for the Gaussian random walk simulation with *η* = 0.1, a dynamic model that precisely matches default EpiFilter. The second column (B, E, H, K) shows results for the sinusoidal simulation, and the third column (C, F, I, L) shows results for the step-change simulation. In panels (D, E, F), vertical dotted lines indicate default parameter values and the vertical solid line indicates both the default and true parameter value. In panels (G-L), solid coloured lines show central estimates (posterior means) and shading shows 95% credible intervals. Black lines (in *R*_*t*_ estimates) and black dots (in predictive *C*_*t*_ estimates) show the true values of *R*_*t*_ and *C*_*t*_ respectively. Predictive coverage of the 95% credible intervals (closer to 95% is better) and the CRPS (lower is better) are shown as text within each figure.

Using these default parameters results in credible intervals for *R*_*t*_ that typically undercover the true value. This is more noticeable for EpiEstim, where coverage of the 95% credible intervals for *R*_*t*_ in the default model ranges from just 8.9% in the sinusoidal simulation, to 74.4% in the step-change simulation. Marginalising out *k* dramatically improves coverage in these models to 81.1% and 94.4% respectively. Default EpiFilter is generally more robust, partially as a result of the default *η* = 0.1 being less extreme with respect to the posterior distribution of *η*, although marginalising this parameter still improves coverage of *R*_*t*_ from 81.1% to 92.2% in the sinusoidal model.

The one-step-ahead predictive coverage of reported cases is also improved by marginalising the smoothing parameter. This is true for all models and simulations considered, but the effect is more pronounced in EpiEstim.

Finally, marginalising the smoothing parameter generally improves (decreases) the CRPS, suggesting that marginalised models produce more accurate predictive distributions of cases than default models. The sole exception is EpiFilter in the step-change simulation, where the CRPS worsens (increases) slightly, although we show in supplementary section 5 that, on average, marginalisation also improves EpiFilter when fit to step-change simulations. The higher CRPS in this specific simulation suggests that the narrower credible intervals produced by default EpiFilter may trade off favourably against its lower predictive coverage of cases. We observe a similar effect in the sinusoidal simulation when comparing EpiEstim and EpiFilter, where the CRPS is slightly lower in EpiEstim (indicating it as the better model), despite EpiFilter having better coverage of predictive cases.

These results depend on the simulated ground truth, and we consider different simulations in supplementary section 5, revealing how the appropriate level of smoothing depends on the underlying epidemic dynamics. Marginalisation becomes more important as the standard deviation of the simulated random walk increases, the frequency of the sinusoidal curve increases, or the step-change becomes more frequent. In all these scenarios, the true *R*_*t*_ is more dynamic and the default smoothing choices are progressively worse at adapting to these changes.

We also find, contrary to intuition, that fitting to greater numbers of daily cases does not necessarily improve inference quality. Supplementary figure S4 demonstrates that, while marginalised EpiFilter is largely robust to sample size, EpiEstim’s coverage worsens as sample size increases in both the default and marginalised models. This occurs as guaranteed misspecification (*R*_*t*_ cannot be constant on [*t* − *k* + 1, *t*] and then constant at a different value on [*t* − *k* + 2, *t* + 2]) results in the model becoming more confident in the incorrect estimate. For similar reasons, if observation noise is large, both models are also expected to degrade as sample size increases.

### The COVID-19 pandemic in New Zealand, August-December 2021

After largely containing the spread of COVID-19, in August 2021 an outbreak of the delta-variant was detected in Auckland, New Zealand, triggering strict non-pharmaceutical interventions. The outbreak featured an initial peak in late August, followed by a subsequent period of decline, and then a second peak in mid-November (figure 3). *R*_*t*_ was repeatedly cited during decision-making, including by the Prime Minister and the Director-General of Health [3, 32, 33, 34, 4, 35, 36].

**Figure 3:**
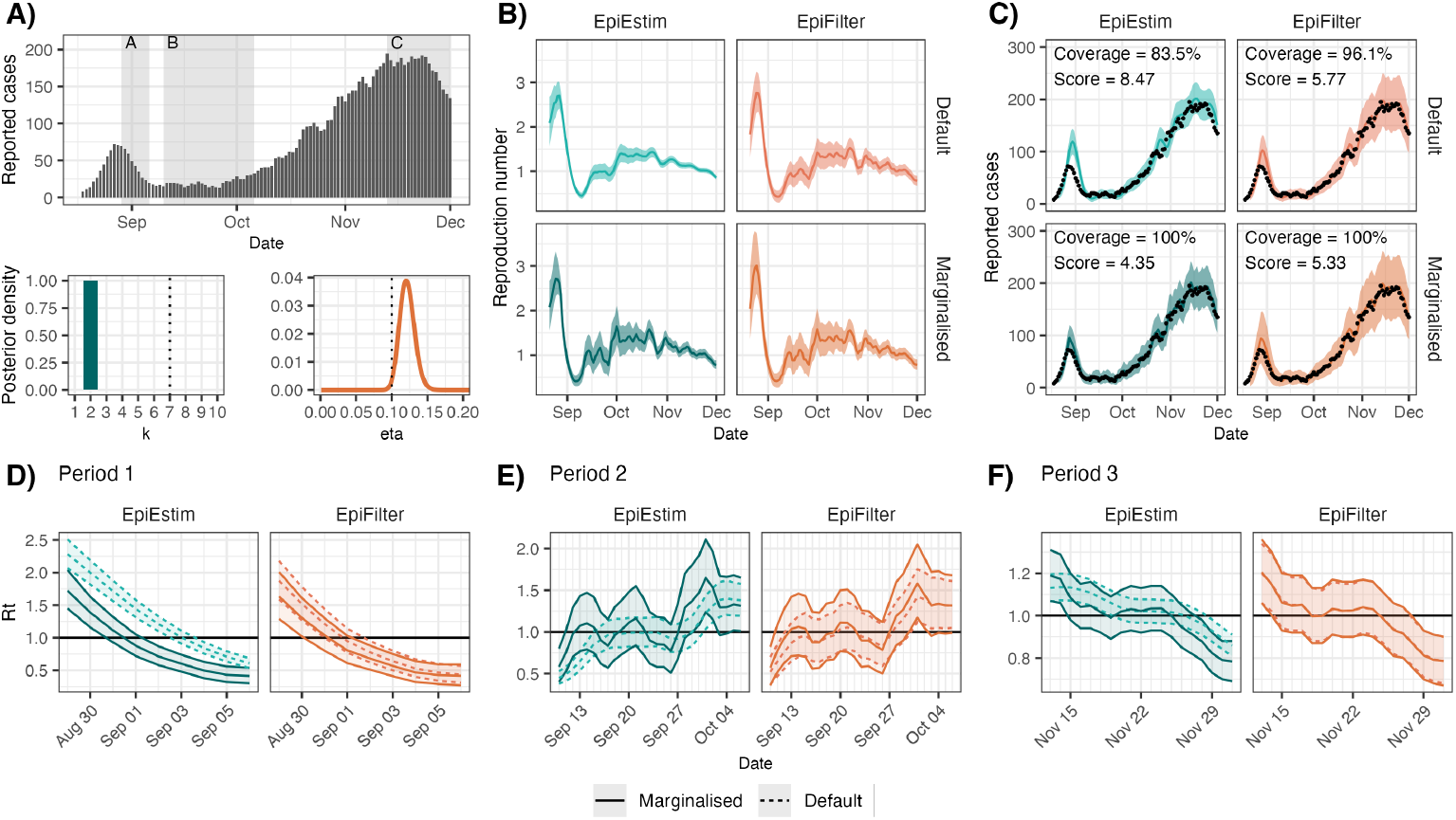
Reported case data and parameter posterior estimates (A), reproduction number estimates (B), predictive cases (C) and reproduction number estimates for selected periods (D, E, F) from fitting the four models to reported case data from the outbreak of the delta-variant of SARS-CoV-2 in Auckland, New Zealand between 11 August 2021 and 1 December 2021. In panel A, vertical dashed lines indicate default parameter values while coloured curves show parameter posterior distributions. In panels (B, C), solid lines show central estimates (posterior means) and shading shows posterior 95% credible intervals. Black points show observed reported cases. In panels (D, E, F), middle lines show central estimates (posterior means) while shading and outer lines show posterior 95% credible intervals. Results for default and marginalised EpiFilter in period 3 (Panel F) are nearly identical and overlap considerably. Our methods improve predictive coverage of EpiEstim and the CRPS of both models.

We fit all four models to reported cases (smoothed using a 5-day moving average to decrease reporting noise; supplementary section 6 tests sensitivity to this assumption) between 11 August and 1 December 2021 (figure 3). Both EpiEstim and EpiFilter exhibit improved CRPS after marginalisation, and EpiEstim exhibits improved predictive coverage after marginalisation. As *R*_*t*_ is unknown, we cannot evaluate its calibration.

As in the simulations, default models oversmooth relative to marginalised models. This is most notable for EpiEstim where almost all posterior mass is on *k* = 2. For EpiFilter, the MAP value of *η* was 0.12 with a 95% credible interval of (0.102, 0.143), which nearly contains the default value. In this case, default EpiFilter is better calibrated than default EpiEstim. However, table 1 still highlights some delays in detection of epidemic growth or decline.

**Table 1:** Dates of the first detection of growth or decline (defined as the upper bound on the 95% credible interval of *R*_*t*_ crossing 1 for growth, or the lower bound crossing 1 for decline), the date that the central estimates of *R*_*t*_ first crossed 1, and the date that the models are first confident in growth or decline (defined as the lower bound on the 95% credible interval of *R*_*t*_ crossing 1 for growth, or the upper bound crossing 1 for decline) for the COVID-19 outbreak in Auckland, New Zealand that started in August 2021. The number of days after the first detection is shown in parentheses, highlighting the substantial delay often observed in default models.

|  | EpiEstim |  | EpiFilter |  |
| --- | --- | --- | --- | --- |
|  | Default | Marginalised | Default | Marginalised |
| Period 1 |  |  |  |  |
| First detection | 3 Sep (+3) | 31 Aug (+0) | 1 Sep (+1) | 31 Aug (+0) |
| Expected $R_t < 1$ | 4 Sep (+3) | 1 Sep (+0) | 1 Sep (+0) | 1 Sep (+0) |
| Confidence in $R_t < 1$ | 4 Sep (+2) | 2 Sep (+0) | 2 Sep (+0) | 2 Sep (+0) |
| Period 2 |  |  |  |  |
| First detection | 15 Sep (+4) | 11 Sep (+0) | 13 Sep (+2) | 11 Sep (+0) |
| Expected $R_t > 1$ | 30 Sep (+17) | 13 Sep (+0) | 20 Sep (+7) | 13 Sep (+0) |
| Confidence in $R_t > 1$ | 1 Oct (+1) | 30 Sep (+0) | 1 Oct (+1) | 30 Sep (+0) |
| Period 3 |  |  |  |  |
| First detection | 20 Nov (+5) | 15 Nov (+0) | 15 Nov (+0) | 15 Nov (+0) |
| Expected $R_t < 1$ | 27 Nov (+9) | 19 Nov (+1) | 19 Nov (+1) | 18 Nov (+0) |
| Confidence in $R_t < 1$ | 29 Nov (+1) | 28 Nov (+0) | 28 Nov (+0) | 28 Nov (+0) |

On 31 August, marginalised EpiFilter first signalled *R*_*t*_ *<* 1. Among the four models, default EpiEstim signalled this change last (3 days later). Notably, both marginalised models were confident that *R*_*t*_ *<* 1 by 2 September, before the possibility of decline had been detected by default EpiEstim. This was a period characterised by daily press conferences and strict stay-at-home orders, estimated to have cost the city NZ$56m per day [37].

On 13 September, the Government announced that because restrictions had *“*…*reduced that value [R*_*t*_*] down to consistently below one”*, they would be relaxed the following week on 21 September [34]. There was considerable interest in *R*_*t*_ over that week, as a resurgence could have triggered prolonged restrictions. This announcement occurred after the first detection of a potential resurgence by the marginalised models (11 September), while default EpiFilter first detected this resurgence on the day of the announcement, and default EpiEstim two days later. The marginalised models first produced central estimates of *R*_*t*_ *>* 1 on the same day as the announcement, while only 7 days and 17 days later that default EpiFilter and default EpiEstim estimate *R*_*t*_ *>* 1, respectively.

Daily reported cases continued to increase until mid-November, before appearing to plateau. While the marginalised models (and default EpiFilter) became uncertain about continued epidemic growth around 15 November, default EpiEstim was still confident that *R*_*t*_ *>* 1 until 20 November (for 5 days longer), a clear example of oversmoothed models being overconfident.

We test our methods on additional real-world datasets in supplementary section 6. These examples highlight that real-world data feature many unknowns with no known ground truth. In every case but one, marginalisation decreased the CRPS, emphasising that the most robust models are those that are able to adapt to these unknowns, as well as the importance of validation on observable quantities.

## Discussion

We have derived and analysed novel likelihoods for two popular *R*_*t*_ estimators. We used these likelihoods to develop posterior distributions for the corresponding smoothing parameters, and marginalised out this parametric uncertainty from estimates of *R*_*t*_. Our algorithms generally improve uncertainty quantification for *R*_*t*_, allowing increased confidence in the calibration of reported credible intervals.

Robust uncertainty quantification is crucial for realworld decision-making. Our methods provide one of the first principled and computationally efficient ways of ensuring the robustness of reported uncertainty in two popular models: EpiEstim [11] and EpiFilter [12]. Furthermore, our methods can be applied to other sequential models (i.e., any model that estimates *R*_*t*_ using *C*_1:*t*_ and then *R*_*t*+1_ using *C*_1:*t*+1_ and so on), such as [20, 38, 39], all of which currently assume fixed values of smoothing parameters.

In addition to real-time decision-making, estimates of *R*_*t*_ are also used in models investigating the impact of non-pharmaceutical interventions [40, 41, 42, 43, 44, 45, 46, 47, 48], the effect of climate [49], or the relationship between mobility and transmission [50]. These models all use EpiEstim to estimate *R*_*t*_, most using *k* = 7. While some correct for smoothing-induced delays by shifting estimates by *k/*2 days, this deterministic correction still ignores uncertainty about *k*. The Bayesian nature of our methods allows for the propagation of uncertainty in *k* through to estimates of *R*_*t*_ and thus to downstream models, such as the consensus estimates of *R*_*t*_ produced by SPI-M in England during the COVID-19 pandemic.

While we focus on parametric uncertainty, by fitting our models to simulated data from a range of dynamic models, our work reveals that marginalising smoothing parameters improves model robustness, even when ignoring structural uncertainty. This is observed by improved coverage of predictive cases and decreased CRPS in simulations and on real data, by improved coverage of *R*_*t*_ in simulations, and is a key finding. As *R*_*t*_ is always unknown, it is not possible to compare coverage of this quantity on real data, and we rely on the CRPS on observables as a proxy for model performance (justified by our results on simulated data).

We used two metrics to evaluate model performance: coverage of the 95% credible intervals and CRPS. The former measures calibration only, while the latter also factors in precision (and simultaneously considers all credible intervals, not just the 95% level). Calibration and precision are often a trade-off: improved calibration can be obtained by decreasing precision. The CRPS is a principled way of balancing these two goals. Alternative scoring rules may be appropriate depending on context [51]. The documentation of [52] summarises scoring rules in an epidemiological context.

While our results suggest that improved CRPS implies improved estimates of *R*_*t*_ even when the model is misspecified, there is no guarantee of performance in such situations. Since misspecification is inevitable when modelling infectious diseases, any scoring rule should be interpreted with caution. One example faced by almost all *R*_*t*_ estimators is serial interval misspecification, addressed in supplementary section 4. Like existing literature [18, 26, 53], we find that misspecified serial interval distributions lead to biased estimates of *R*_*t*_, although estimates near *R*_*t*_ = 1 are more robust. Another example of model misspecification comes from observation noise: EpiEstim and EpiFilter assume that reported cases follow the Poisson renewal model and that the appropriate level of smoothing is fixed over time (supplementary section 7). We demonstrate that our methods help with robustness to observation noise in supplementary section 5.3, but the lack of explicitly representing such noise remains a limitation of both models. We chose the New Zealand dataset as an example of high-quality data with limited reporting biases to reduce the impact of this limitation.

Alternative parameter selection procedures for EpiEstim have previously been proposed. In the supplement of [11], the authors suggest selecting *k* such that the window contains sufficient cases to reduce the posterior coefficient-of-variation to a desired level. Depending on philosophy, this is either a subjective decision about the bias-variance trade-off, or a way of choosing parameters to obtain a desired confidence level. In either case, choosing a value of *k* with low likelihood will lead to poor model calibration. Alternatively, Parag and Donnelly [25] proposed an information-theoretic approach to selecting *k* called APEestim. While their approach is principled, it results in the selection of *k* shifted by one unit compared to the MAP value from our method, and does not allow for the marginalisation of uncertainty when there are multiple plausible values of *k* (supplementary section 8). EpiEstim and EpiFilter are Bayesian estimators, thus marginalising smoothing parameters is a more justified approach than selecting a single value. This is supported by supplementary section 2, where two implementations of a different *R*_*t*_ estimator (EpiLPS [22]) are compared: one implementation optimises the smoothing parameters while the other performs marginalisation. We find marginalisation continues to offer improved performance and robustness.

Other methods approach the smoothing problem differently. For example, EpiNow2 [19] models *R*_*t*_ using a Gaussian process. The smoothness of this model is determined by the covariance kernel, which is estimated alongside *R*_*t*_. Comparisons with additional methods (EpiNow2 [19], EpiLPS [22], and rtestim [26]) are included in supplementary section These methods include features such as explicitly representing observation noise, at the cost of increased mathematical complexity and decreased interpretability, when compared to EpiEstim and EpiFilter. We also provide novel methodology for finding posterior predictive distributions and calculating CRPS values for these models, providing model comparison techniques using observed data. While EpiEstim is often outperformed by these methods, there is value in the simplicity and interpretability of a sliding window. Understanding the nuances of various approaches to smoothing and hence inference-based decision-making will form a future study.

The August 2021 outbreak of SARS-CoV-2 in New Zealand provides a pertinent example of the practical importance of smoothing assumptions. *R*_*t*_ was first reported by officials as being less than 1 on 29 August, two days before any of our models. While official models were also based on the renewal model, among other differences (e.g. accounting for asymptomatic infections), they approached the smoothing problem differently, assuming *R*_*t*_ was fixed prior to the lockdown on 18 August, and then step-changed to a different fixed value after the lockdown [31]. These piecewise-constant assumptions allowed more data to inform each estimate of *R*_*t*_, reducing uncertainty. However, if these assumptions were incorrect then uncertainty about *R*_*t*_ will have been underestimated.

It is often argued that public health decision-making should be “data-driven”, with *R*_*t*_ frequently featuring as an example of such “data” [54]. However, without an accurate representation of uncertainty, estimates of *R*_*t*_ risk being influenced more by assumptions than the underlying data. As demonstrated on both simulated and real-world data, public health decisions made using oversmoothed estimates of *R*_*t*_ will be delayed and overconfident relative to decisions made using estimates with more robust uncertainty quantification. Fortunately, our methods provide a simple and computationally efficient way to improve the robustness of these estimates, and to benchmark the uncertainty surrounding smoothing assumptions.

## Additional material

In addition to the supplementary material, all code is available on the GitHub repository. This repository contains all code necessary to reproduce the results in this paper, as well as Julia implementations of both EpiEstim and EpiFilter. Tutorials for the use of these methods are provided on the corresponding website.

## Acknowledgements

N.S. acknowledges support from the Oxford-Radcliffe Scholarship from University College, Oxford, the EPSRC CDT in Modern Statistics and Statistical Machine Learning (Imperial College London and University of Oxford), and A. Maslov for studentship support. K.V.P. acknowledges funding from the MRC Centre for Global Infectious Disease Analysis (Reference No. MR/X020258/1) funded by the UK Medical Research Council. This UK-funded grant is carried out in the frame of the Global Health EDCTP3 Joint Undertaking. The funders had no role in study design, data collection and analysis, decision to publish, or manuscript preparation.

## Data availability statement

All data used in this study is publicly available from [28]. All data and code used in our analysis is available at https://github.com/nicsteyn2/RobustRtEstimators.

## Footnotes

1 Technically, the terms EpiEstim and EpiFilter refer to the associated software packages. We follow popular convention and use these terms to describe the method.

