## Supplementary text for "Robust uncertainty quantification in popular estimators of the instantaneous reproduction number"

**7 Stepwise likelihoods** **31**

**8 APE comparison** **32**

**9 Testing grid-sizes** **34**

### 1 Derivations

#### 1.1 General notes

Before considering specific derivations, we highlight some useful tools that are common between estimators.

##### 1.1.1 Predictive decomposition of the likelihood

Letting $\theta$ be the parameter(s) of interest (typically $k$ for EpiEstim or $\eta$ for EpiFilter), the log-likelihood of $\theta$ at time-step $t$ is defined as:

$$\ell(\theta|C_{1:t}) = \log P(C_{1:t}|\theta) \tag{S1}$$

This can be decomposed into the sum of the one-step-ahead predictive log-likelihoods:

$$\ell(\theta|C_{1:t}) = \sum_{s=1}^{t} \log P(C_s|C_{1:s-1}, \theta) \tag{S2}$$

That is, the likelihood of observing all data $C_{1:t}$ is equal to the product of the likelihoods of observing each data point $C_s$ given the data up to that point $C_{1:s-1}$.

##### 1.1.2 Grid-based approximations

We frequently leverage grid-based approximations to various distributions in EpiFilter, and to a lesser extent in EpiEstim. These are necessary (in some cases) when considering three specific variables: the reproduction number $R_t$, EpiFilter's smoothing $\eta$, and predictive cases $C_{t+1}$.

We denote the grid of values for $R_t$ using $\mathcal{R}$, typically using one of length $|\mathcal{R}| = 1000$:

$$\mathcal{R} = \{0.01, 0.02, \ldots, 10.000\}$$

We denote the grid of values for $\eta$ using $\mathcal{E}$, typically using one of length $|\mathcal{E}| = 1000$:

$$\mathcal{E} = \{0.001, 0.002, \ldots, 1.000\}$$

Finally, we denote the grid of values for predictive cases $C_{t+1}$ using $\mathcal{C}$, and typically use:

$$\mathcal{C} = \{0, 1, 2, \ldots, 10 \max C_{1:T}\}$$

where $10 \max C_{1:T}$ (where $C_{1:T}$ is the observed cases series) is chosen to be sufficiently large to capture the majority of the predictive distribution. This is checked by ensuring that the sum of the predictive distribution over $\mathcal{C}$ is close to 1.

A grid-based approximation to a distribution for $R_t$ (for example) is then given by the set of values $\{P(R_t = r| \cdots)\}_{r \in \mathcal{R}}$.

We test the sensitivity of our results to the choice of grids in supplementary material section 9.

##### 1.1.3 Wind-in periods

Wind-in periods may be necessary for two reasons. Firstly, if a model is initialised mid-outbreak, then reported cases at time $t$ (for low $t$) depend on reported cases prior to the initialisation of the model. A wind-in period is thus necessary to ensure a sufficient number of past data points are available to inform the model, the necessary duration of which depends on the specific application. Secondly, the derivation of a one-step-ahead likelihood requires the predictive distribution of $R_{t+1}$, which is derived from the posterior distribution of $R_t$, which requires at least one day of prior data. Thus, model likelihoods can only be calculated from time-step $t = 3$ onwards.

In the 2021 August COVID-19 outbreak in Auckland, New Zealand, the model is initialised alongside the first reported case, thus only a 3-day wind-in period is necessary. For simulated data, we assume that the model was initialised mid-outbreak, and thus use a 10-day wind-in period.

For simulated results, we also normalise the generation time distribution to sum to 1 over $[1, t]$, reducing the impact of a wind-in period that does not cover the entire support of the generation time distribution.

#### 1.2 EpiEstim

##### 1.2.1 Model description

EpiEstim assumes that $R_t$ is fixed over the interval $[t-k+1, t]$. The likelihood of observing $C_{t-k+1:t}$ given $R_t$, $k$, and $C_{1:t-k}$ is then given by the product of the Poisson renewal model likelihood (equation 1) for each day $s$ in the interval $[t-k+1, t]$:

$$P(C_{t-k+1:t}|R_t, C_{1:t-k}, k) = \prod_{s=t-k+1}^{t} P(C_s|R_t, \Lambda_s) = \prod_{s=t-k+1}^{t} \frac{(R_t\Lambda_s)^{C_s} e^{-R_t\Lambda_s}}{C_s!} \tag{S3}$$

A conjugate (shape-rate-parameterised) Gamma$(\alpha, \beta)$ prior distribution is assumed for $R_t$, thus the posterior distribution for $R_t$ given $k$ and $C_{1:t}$ is also Gamma-distributed:

$$\begin{aligned} P(R_t|C_{1:t}, k) &\propto P(C_{t-k+1:t}|R_t, C_{1:t-k}, k)P(R_t) \\ &\propto \left(R_t^{\sum C_s} e^{-R_t \sum \Lambda_s}\right)\left(R_t^{\alpha-1} e^{-\beta R_t}\right) \\ &= R_t^{\alpha+\sum C_s - 1} e^{-R_t(\beta + \sum \Lambda_s)} \\ &\sim \text{Gamma}\left(\alpha_{t,k} = \alpha + \sum_{s=t-k+1}^{t} C_s, \quad \beta_{t,k} = \beta + \sum_{s=t-k+1}^{t} \Lambda_s\right) \end{aligned} \tag{S4}$$

The shape parameter $\alpha_{t,k}$ at time $t$ is the shape parameter of the prior distribution $\alpha$ plus the sum of the previous $k$ days' case counts. The rate parameter $\beta_{t,k}$ at time $t$ is the rate parameter $\beta$ of the prior distribution plus the sum of the previous $k$ days' force of infection[1]. We make regular use of $\alpha_{t,k}$ and $\beta_{t,k}$ in the following derivations.

[1] Through this lens, EpiEstim's prior parameters $\alpha$ and $\beta$ can be viewed as the assumed number of observed infections and force-of-infection before any data are collected.

##### 1.2.2 Likelihood for $k$

We use the predictive decomposition of the likelihood (equation S2) to derive the likelihood of $k$ given $C_{1:t}$. First note that:

$$P(C_s|C_{1:s-1},k) = \int P(C_s|R_s,C_{1:s-1},k)P(R_s|C_{1:s-1},k)dR_s \tag{S5}$$

The first term in the integral is the Poisson renewal model likelihood (equation 1), and the second term is the predictive distribution for $R_s$ given $C_{1:s-1}$ and $k$. We find this predictive distribution now.

Applying Bayes' theorem, leveraging EpiEstim's assumed independence of $R_s$ from $C_{1:s-k}$, and substituting in the known distributions gives:

$$\begin{aligned}
P(R_s|C_{1:s-1},k) &\propto P(C_{t-k+1:s-1}|R_s,C_{1:s-k},k)P(R_s) \\
&= P(R_s)\prod_{u=s-k+1}^{s-1} P(C_u|R_s,C_{1:u-1},k) \\
&\propto R_s^{\alpha-1}e^{-\beta R_s}\prod_{u=s-k+1}^{s-1} R_s^{C_u}e^{-R_s\Lambda_s} \\
&= R_s^{\alpha+\sum_{u=s-k+1}^{s-1}C_u-1}e^{-R_s\left(\beta+\sum_{u=s-k+1}^{s-1}\Lambda_u\right)}
\end{aligned} \tag{S6}$$

Thus, the predictive distribution for $R_s$ given $C_{1:s-1}$ and $k$ is Gamma-distributed with shape parameter $\alpha_{s-1,k-1}$ and rate parameter $\beta_{s-1,k-1}$ (where $\alpha_{s-1,k-1}$ and $\beta_{s-1,k-1}$ are once again defined in equations 2).

This is not a surprising result. By (EpiEstim's) definition, $R_s$ is only informed by data between time-steps $s-k+1$ and $s$, however the predictive distribution for $R_s$ must not include $C_s$, so the parameters of the posterior distribution for $R_s$ are the sum over $k-1$ days of data. This is the key difference between our approach to "selecting" $k$ and that advocated by the accumulated prediction error (APE) approach of [8] (supplementary section 8), where the APE approach temporarily assumes that $R_s$ is fixed for the preceding $k+1$ days, so that the predictive distribution can be informed by $k$-days of data.

The predictive distribution for $C_s$ conditional on $C_{1:s-1}$ and $k$ is a Poisson-Gamma mixture, and thus is negative binomial:

$$C_s|C_{1:s-1},k \sim \text{NegativeBinomial}\left(r=\alpha_{s-1,k-1}, p=\frac{\beta_{s-1,k-1}}{\Lambda_s+\beta_{s-1,k-1}}\right) \tag{S7}$$

Finally, the log-likelihood of $k$ given $C_{1:t}$ is given by the sum of the log-likelihoods of each day $s$ in the interval $[t-k+1,t]$:

$$\ell(k|C_{1:t}) = \sum_{s=1}^{t}\log\text{NegBinPMF}\left(C_s|r=a_{s-1,k-1}, p=\frac{\beta_{s-1,k-1}}{\Lambda_s+\beta_{s-1,k-1}}\right) \tag{S8}$$

##### 1.2.3 Posterior distribution for $k$

The log-likelihood $\ell(k|C_{1:t})$ (equation S8) is sufficient for calculating quantities such as the maximum likelihood estimate of $k$. However, in order to marginalise over $k$, we require the posterior distribution for $k$ given $C_{1:t}$.

We typically use a Uniform prior distribution over a sufficiently wide range of $k$ values, say $k \in \{1, 2, \ldots, 30\}$, although other discrete prior distributions are possible. The posterior distribution for $k$ is then proportional to the exponential of $\ell(k|C1 : t)$:

$$P(k|C_{1:t}) = \begin{cases} c \exp\left(\ell(k|C_{1:t})\right) & k \in \{1, 2, \ldots, 30\} \\ 0 & \text{otherwise} \end{cases} \tag{S9}$$

where the normalising constant $c$ is chosen such that $\sum_{k=1}^{30} P(k|C_{1:t}) = 1$. We note here that this posterior distribution is exact and does not rely upon any approximation.

##### 1.2.4 Marginalising over $k$

To present estimates of $R_t$ that do not depend on our choice of $k$, we marginalise over $k$ using the posterior distribution derived above (equation S9):

$$P(R_t|C_{1:t}) = \sum_k P(R_t|C_{1:t}, k)P(k|C_{1:t}) \tag{S10}$$

The first term in the summation is given by equation S4 and the second term is given by equation S9.

We typically calculate this over a grid $\mathcal{R}$ of $R_t$ values as defined in supplementary material section 1.1.2. The resulting posterior mean and credible intervals for $R_t$ are then calculated from this grid and are statements about $R_t$ given the data $C_{1:t}$ (and implicitly the model structure), but independent of any specific choice of $k$.

##### 1.2.5 Posterior predictive distribution

We previously derived the predictive distribution for $C_t$ given $C_{1:t-1}$, conditional on some value of $k$ (equation S7). We can also marginalise over $k$ to obtain the predictive distribution for $C_t$ given $C_{1:t-1}$:

$$P(C_t|C_{1:t-1}) = \sum_k P(C_t|C_{1:t-1}, k)P(k|C_{1:t-1}) \tag{S11}$$

The first term in the summation is given by equation S7 and the second term is given by equation S9.

We typically calculate this over a grid $\mathcal{C}$ of potential $C_t$ values as defined in supplementary material section 1.1.2.

###### 1.2.6 Practical notes

At time-step $s = 1$, there is no past data to inform the predictive distribution for $R_s$ (equation S6). Therefore, $R_1$ is independent of $k$ and provides no information about the posterior distribution for $k$. When calculating these distributions, we typically begin related summations at $s = 2$.

#### 1.3 EpiFilter

##### 1.3.1 Model description

EpiFilter models the evolution of $R_t$ using a Gaussian random walk with standard deviation $\eta\sqrt{R_{t-1}}$:

$$R_t|R_{t-1} \sim \text{Normal}\left(R_{t-1}, \eta\sqrt{R_{t-1}}\right) \tag{S12}$$

Given the posterior distribution for $R_{t-1}$ at time-step $t-1$, the posterior distribution for $R_t$ is found by applying the Bayesian filtering equations. Specifically, we first find the update distribution:

$$P(R_t|C_{1:t-1}, \eta) = \int P(R_t|R_{t-1}, \eta)P(R_{t-1}|C_{1:t-1}, \eta)\ dR_{t-1} \tag{S13}$$

and then use the Poisson renewal likelihood to find the filtering distribution:

$$P(R_t|C_{1:t}, \eta) \propto P(C_t|R_t, C_{1:t-1})P(R_t|C_{1:t-1}, \eta) \tag{S14}$$

Analytical forms for these distributions are not readily available, so they are approximated using a grid of $R_t$ values $\mathcal{R}$. This replaces the integral in equation S13 with a summation over $R_{t-1} \in \mathcal{R}$, and allows us to normalise the filtering distribution in equation S14 by summing over the grid.

##### 1.3.2 Likelihood for $\eta$

We use a similar approach to EpiEstim to derive the likelihood of $\eta$ given $C_{1:t}$. Re-writing equation S5 in terms of EpiFilter's $\eta$:

$$P(R_s|C_{1:s-1}, \eta) = \int P(C_s|R_s, C_{1:s-1}, \eta)P(R_s|C_{1:s-1}, \eta)dR_s \tag{S15}$$

The first term in the integral is the Poisson renewal model likelihood (equation 1), and the second term is the update distribution for $R_s$ given $C_{1:s-1}$ and $\eta$, which is a by-product from solving the filtering equations (specifically, equation S13). Leveraging the grid-based approximation, we write this as:

$$P(R_s|C_{1:s-1}, \eta) = \sum_{r \in \mathcal{R}} P(R_s|R_{s-1} = r, \eta)P(R_{s-1} = r|C_{1:s-1}, \eta) \tag{S16}$$

Then the probability of observing $C_s$ given $C_{1:s-1}$ and $\eta$ is found by marginalising over $R_s$:

$$P(C_s|C_{1:s-1},\eta) = \sum_{r\in\mathcal{R}} P(C_s|R_s = r, C_{1:s-1})P(R_s = r|C_{1:s-1},\eta) \tag{S17}$$

Thus, the log-likelihood of $\eta$ at time-step $t$ can be written as:

$$\ell(\eta|C_{1:t}) = \sum_{s=1}^{t} \log\left(\sum_{r\in\mathcal{R}} P(C_s|R_s = r, C_{1:s-1})\left[\sum_{r'\in\mathcal{R}} P(R_s = r|R_{s-1} = r',\eta)P(R_{s-1} = r'|C_{1:s-1},\eta)\right]\right) \tag{S18}$$

##### 1.3.3 Posterior distribution for $\eta$

The log-likelihood $\ell(\eta|C_{1:t})$ (equation S18) is sufficient for calculating quantities such as the maximum likelihood estimate of $\eta$. However, in order to marginalise over $\eta$, we require the posterior distribution for $\eta$ given $C_{1:t}$.

We typically use a Uniform prior distribution over a sufficiently wide range of $\eta$ values, say $\eta \in (0, 0.5)$. The posterior distribution for $\eta$ is then proportional to the exponential of $\ell(\eta|C1 : t)$:

$$P(\eta|C_{1:t}) = \begin{cases} c\exp\left(\ell(\eta|C_{1:t})\right) & \eta \in (0, 0.5) \\ 0 & \text{otherwise} \end{cases} \tag{S19}$$

As no analytical form for this distribution is available, we use another grid-approximation (this time over $\eta \in \mathcal{E}$), chosen such that $\sum_{\eta\in\mathcal{E}} P(\eta|C_{1:t}) = 1$. This posterior distribution is not exact in the same sense as the equivalent distribution for $k$ in EpiEstim, but we show that it is a good approximation for sufficiently large $|\mathcal{E}|$ (supplementary material section 1.1.2).

##### 1.3.4 Marginalising over $\eta$

To present estimates of $R_t$ that do not depend on our choice of $\eta$, we marginalise over $\eta$ using the posterior distribution derived above (equation S19):

$$P(R_t|C_{1:t}) = \sum_{\eta\in\mathcal{E}} P(R_t|C_{1:t},\eta)P(\eta|C_{1:t}) \tag{S20}$$

The first term in the summation is given by equation S14 and the second term is given by equation S19. We typically calculate this over a grid $\mathcal{R}$ of $R_t$ values.

##### 1.3.5 Posterior predictive distribution

We marginalise over $\eta$ in equation S17 to find the marginal posterior predictive distribution:

$$P(C_t|C_{1:t-1}) = \sum_{\eta\in\mathcal{E}} P(C_t|C_{1:t-1},\eta)P(\eta|C_{1:t-1}) \tag{S21}$$

The first term in the summation is given by equation S17 and the second term is given by equation S19.

###### 1.3.6 Marginal smoothing posterior distribution

We focus on real-time analysis in this paper, and thus focus on EpiFilter's *filtering distribution*, the posterior distribution for $R_t$ given data up-to time $t$. The full implementation of EpiFilter also includes a *smoothing distribution* that incorporates both past and future data in estimates of $R_t$. It is easy to marginalise out $\eta$ from this smoothing distribution:

$$P(R_t|C_{1:T}) = \int P(R_t|C_{1:T}, \eta)P(\eta|C_{1:T})\ d\eta \tag{S22}$$

where $P(\eta|C_{1:T})$ is simply the posterior distribution for $\eta$ at the final time-step $T$. That is, the predictive decomposition of the likelihood produces the correct likelihood for $\eta$ regardless of whether we are considering the filtering or smoothing distribution.

This marginal smoothing posterior distribution is used and compared with other methods in supplementary material section 2.

##### 1.4 Continuous ranked probability score

The continuous ranked probability score (CRPS) is a measure of the accuracy of a probabilistic forecast. For a given time-step $t$, the CRPS is defined as:

$$CRPS(F_t, C_t) = \int (F_t(y) - \mathbb{I}(y \geq C_t))^2 dy \tag{S23}$$

where $F_t(y)$ is the predictive distribution for $C_t$ at time $t$, and $\mathbb{I}(y \geq C_t)$ is the indicator function that is 1 if $y \geq C_t$ and 0 otherwise. The CRPS is a measure of the distance between the predictive distribution and the observed value. The CRPS for a model is the average CRPS over all time-steps in the dataset, with lower scores representing better forecasts.

The CRPS is a strictly proper scoring rule, meaning that the optimal forecast is the true distribution. It is also a strictly consistent scoring rule, meaning that the optimal forecast converges to the true distribution as the number of observations increases.

We calculate the CRPS using the following formula:

$$CRPS(F_t, C_t) = \sum_{y \in \mathcal{C}} F_t(y) - \mathbb{I}(y \geq C_t) \tag{S24}$$

where $F_t(y)$ is the CDF for the predictive distribution of $C_t$ (found either analytically in the case of default EpiEstim, or by taking the cumulative sum over the grid-approximation in the case of marginalised EpiEstim and EpiFilter).

CRPS should only be compared between models that have been fit to the same data, as it is a measure of the accuracy of a forecast, not the quality of a model. In supplementary section 5 we average results over multiple simulated epidemics. To ensure that CRPS values are comparable, in this section, we report the average of CRPS scores relative to the default EpiEstim model.

### 2 Smoothing posterior distributions and comparisons with additional methods

*The material in this supplementary section has been published as a live document online. We provide a copy here for record, although the reader may find the online document more up-to-date.*

#### 2.1 Background

Many state-of-the-art $R_t$ estimation methods target the smoothing distribution, rather than the filtering distribution (see below). Examples include the smoothing version of EpiFilter [6], EpiNow2 [1], EpiLPS [3], and rtestim [4]. We compare and contrast these methods on simulated data here.

For each method, we:

- Find the posterior distribution for $R_t$
- Find the posterior predictive distribution for observed cases $\tilde{C}_t$
- Calculate the calibration of the implied 95% credible intervals for $R_t$ and $C_t$
- Calculate the CRPS of the posterior predictive distribution for observed cases

We use $\tilde{C}_t$ to highlight when we are treating reported cases as a random variable, instead of as observed data.

Unlike the other methods listed, rtestim is frequentist in nature. In this case, we replace posterior means/modes and credible intervals with estimates and confidence intervals. All of the above metrics still apply and carry comparable interpretations.

The posterior predictive distribution obtained from smoothing methods typically provides within-sample estimates. That is, the observed value $C_t$ is used to find the posterior predictive distribution of $\tilde{C}_t$. This can occur either indirectly (observed $C_t$ informs $R_t$ estimates, which then are used to predict $\tilde{C}_t$) or directly (observed $C_t$ informs estimates of latent infection incidence, from which $\tilde{C}_t$ are generated). This changes the interpretation of CRPS from a score of future predictions to a score of how well the data-generating mechanism is modelled.

In many cases, existing methodology had to be extended to find the posterior predictive distributions or to calculate the CRPS. Model descriptions are otherwise kept to a minimum. Where possible, default options of these models are used, although minor modifications are made where defaults lead to obviously suboptimal results.

Code to explicitly reproduce these results and figures is provided on GitHub.

##### 2.1.1 Filtering versus smoothing

The **filtering posterior distribution** depends only on past data and is written $P(R_t|C_{1:t})$, whereas the **smoothing posterior distribution** depends on all data and is written $P(R_t|C_{1:T})$ [9].

When producing estimates at the most recent time-step, only the filtering distribution is accessible, as future data are not yet available, whereas the smoothing distribution is generally preferred for retrospective analysis, as it incorporates all available data. Note that, at time $t = T$, the filtering and smoothing distributions are equivalent.

In the main paper, we focus on methods for estimating $R_t$ in real-time. That is, we target the filtering distribution rather than the smoothing distribution. In this supplementary section we consider the latter.

#### 2.2 Methods

##### 2.2.1 EpiFilter (smoothing)

We outline how EpiFilter can be used to find the marginal smoothing posterior distribution in supplementary section 1.3.6.

The posterior smoothing distribution for observed case incidence is found by marginalising over $R_t$ from the smoothing posterior distribution for $R_t$ (instead of the predictive distribution for $R_t$):

$$P(\tilde{C}_t|C_{1:T}) = \int P(\tilde{C}_t|R_t, C_{1:T})P(R_t|C_{1:T})\, dR_t$$

For simplicity, we use the renewal model for $P(\tilde{C}_t|R_t, C_{1:T})$, even though this ignores future reported case incidence. That is, future case data $C_{t:T}$ only features in $P(\tilde{C}_t|C_{1:T})$ via $R_t$.

##### 2.2.2 EpiNow2

Rather than using a Gaussian random walk (as in EpiFilter) or fixed sliding windows (as in EpiEstim), EpiNow2 models the evolution of $R_t$ using a Gaussian process. The default implementation then assumes that latent infection incidence follows a deterministic renewal model. Reported cases are assumed to be negative binomially distributed around the true infection, with a day-of-the-week effect that is estimated during the fitting process. That is, EpiNow2 accounts for both process noise (in the evolution of $R_t$) and observation noise (in the distribution for reported cases).

Smoothness in $R_t$ is primarily controlled by the Gaussian process kernel (default Matern 3/2, with lengthscale $\ell$ and magnitude $\alpha$). In particular, prior assumptions about the lengthscale $\ell$ have a significant impact on the smoothness of resulting estimates. It is also expected that prior assumptions about the observation overdispersion will have a secondary effect on the smoothness of $R_t$ estimates (as this impacts the trade-off between process and observation noise).

The EpiNow2 package provides pre-built functionality to extract central estimates and credible intervals for $R_t$ and observed cases. As far as we are aware, the package does not provide a method to calculate the CRPS, so we provide our own sample-based implementation.

Samples from the posterior distribution for reported cases $\{x_t^{(i)}\}_{i=1}^N$ at time $t$ are extracted from the Stan fit object. We then use the sample-based CRPS estimator:

$$\text{CRPS}_t = \frac{1}{N}\sum_{i=1}^{n} |x_t^{(i)} - C_t| - \frac{1}{2N^2}\sum_{i=1}^{n}\sum_{j=1}^{n} |x_t^{(i)} - x_t^{(j)}|$$

Calculating this for each time-step $t$ and taking the average gives the CRPS for observed case incidence. Code to reproduce this is available on GitHub.

By default, EpiNow2 uses a log normal prior distribution for the lengthscale $\ell$ with mean 21 days and standard deviation 7 days. We also test an alternative and less informative inverse-gamma prior distribution provided with the package, which performs considerably better.

###### 2.2.3 EpiLPS (MAP)

EpiLPS models latent infection incidence using Bayesian P-splines. Given infection incidence $\mu(t)$ and overdispersion parameter $\rho$, reported cases are assumed to be negative binomially distributed around $\mu(t)$. In the maximum a posteriori (MAP) version of EpiLPS, central estimates of $R_t$ are calculated using a plug-in estimate of the posterior mean incidence $\hat{\mu}(t)$ and uncertainty is derived using a delta method. Like EpiNow2, EpiLPS explicitly accounts for both process noise (in the splines used to model infection incidence) and observation noise (in the distribution for reported cases).

Smoothness in $R_t$ is primarily controlled by parameter $\lambda$, where larger values penalise sharp changes in infection incidence. A hierarchical prior distribution is assumed for $\lambda$. Prior assumptions about the overdispersion parameter $\rho$ also likely impact the smoothness of $R_t$ estimates, as this controls how much noise is associated with the observation process rather than the underlying epidemic process. The MAP version of EpiLPS selects optimal values of $\lambda$ and $\rho$ using an optimization routine.

While $K = 30$ is used as the default number of splines, we find that this leads to inaccurate inference on our example datasets. For our examples, we increase this to $K = 100$, which allows for more flexible inference.

We provide two extensions to the EpiLPS package:

1. Methods to sample from the posterior distribution for reported cases.
2. A CRPS estimator for the posterior distribution for reported cases.

Infection incidence is defined as $\mu(t) = \exp(\theta^T b(t))$, where $\theta$ is an estimated vector of spline coefficients and $b(t)$ are the basis functions evaluated at time $t$. The MAP version of EpiLPS uses a multivariate Gaussian approximation to $\theta$ at the MAP estimate of $\lambda$. We sample from this distribution:

$$\theta^{(i)} \sim MVN(\hat{\theta}, Q_\lambda^{-1})$$

and use these samples to generate samples of latent infection incidence:

$$\mu^{(i)}(t) = \exp(\theta^{(i)T} b(t))$$

from which samples of observed data are generated:

$$C_t^{(i)} \sim \text{NegBin}(\mu^{(i)}(t), \rho)$$

where $\rho$ is the MAP value of the overdispersion parameter. This is repeated $N$ times (default $N = 1000$) at each value of $t$. The mean of these $N$ samples is reported as the central estimate, with 95% credible intervals obtained by taking the 2.5th and 97.5th percentile values.

The CRPS is calculated similarly, first by sampling $\mu^{(i)}(t)$ as above and then using the *crps_nbinom()* function from the *scoringRules* package in R to calculate the CRPS value for each sampled $\mu^{(i)}(t)$ and $C_t$. The mean of these $N$ CRPS values is reported as the CRPS for the $t^{th}$ observed case incidence.

Some code is available on GitHub, although copyright limitations prevent us from providing the full implementation, in which case we outline the steps required to reproduce the analysis.

###### 2.2.4 EpiLPS (MALA)

The Metropolis-adjusted Langevin algorithm (MALA) version of EpiLPS replaces the Laplace approximation and optimisation routine with a full MCMC-type sampler. This has the advantage of returning a posterior distribution on $\lambda$ and $\rho$, as well as properly marginalising out uncertainty about these quantities. As the true posterior distribution of spline coefficients $\theta$ is targeted, we also expect the posterior distributions for $R_t$ and observed cases incidence to be more accurate. This comes at a cost of slightly increased computational complexity, although we do not find this to be prohibitive.

The default version of EpiLPS(MALA) does not return the MCMC sampling object. In order to access this, we created a customised version of *estimRmcmc.R* that includes *MCMC=MCMCout* in the *"outputlist"* of the function. This is the sole change required to this script, although running it requires having local copies of some files from the EpiLPS package. A list of the required files is given here.

We make the same two extensions to EpiLPS(MALA) as we did to EpiLPS(MAP):

1. Methods to sample from the posterior distribution for reported cases.
2. A CRPS estimator for the posterior distribution for reported cases.

Samples of reported case incidence are extracted from the MCMC sampling object. For each sample $i = 1, \ldots, N$ and time-step $t = 1, \ldots, T$ we sample:

$$C_t^{(i)} \sim \text{NegBin}(\exp(\theta^{(i)T} b(t)), \rho^{(i)})$$

where $\theta^{(i)}$ and $\rho^{(i)}$ are samples from the MCMC sampling object. Note the use of $\rho^{(i)}$ instead of the MAP value of $\rho$, ensuring we are appropriately accounting for uncertainty in this parameter.

The CRPS is calculated from sampled $C_t^{(i)}$ using the same approach as for EpiNow2, relying upon the sample-based CRPS estimator (see above).

Some code is available on GitHub, although copyright limitations prevent us from providing the full implementation, in which case we outline the steps required to reproduce our analysis.

###### 2.2.5 rtestim

In contrast to the aforementioned Bayesian methods, rtestim is grounded in a frequentist framework. $R_t$ is modelled using piecewise cubic functions with $\ell_1$ regularisation on the divided differences. This regularisation enforces sparsity in changes in $R_t$, allowing for locally adaptive smoothness, a key advantage over the other methods considered here that assume global smoothness.

A tuning parameter $\lambda$ controls the strength of this regularisation, with larger values enforcing smoother estimates of $R_t$. The optimal value of this parameter is automatically chosen by rtestim using cross-validation.

rtestim uses the delta method to calculate confidence intervals. Built-in functions are provided to calculate these for $R_t$ and observed case incidence, for any user-specified significance level. To match the other methods we use a 95% confidence level.

While the frequentist framework of rtestim does not admit a posterior distribution for observed case incidence, we can still use CRPS to measure how well the confidence intervals approximate the observed distribution of the data. To do this, we treat the bounds of the confidence intervals at different significance levels as defining quantiles of an empirical CDF. That is, at each time-step $t$, we find $x_t^{(i)}$ such that $q^{(i)} = F(x_i^{(t)})$ for $q^{(i)} = 0.01, 0.02, \ldots, 0.99$. We then approximate the CRPS numerically at time-step $t$ using the trapezoidal rule:

$$CRPS_t = \sum_{i=1}^{n-1} \frac{1}{2} \left( x_t^{(i+1)} - x_t^{(i)} \right) \left[ \left( q^{(i+1)} - \mathbb{I}(x_t^{(i+1)} \geq y_t) \right)^2 + \left( q^{(i)} - \mathbb{I}(x_t^{(i)} \geq y_t) \right)^2 \right]$$

Averaging $CRPS_t$ over all time-steps $t$ gives the CRPS for observed case incidence. Code to reproduce this is available on GitHub.

###### 2.2.6 Additional methodological notes

Each method handles the wind-in period differently, particularly when $t$ is small compared to generation times. This can become fairly complicated, and is not the point of this supplementary section. When estimating or calculating coverage and CRPS, we use time-steps $t = 10, \ldots, T$ to avoid differences from this period impacting our results.

Posterior parameter values were calculated using a grid-based approach for EpiFilter. For EpiLPS (MALA) and EpiNow2, a sampling approach was required. The mean, 2.5th and 97.5th quantiles of these samples were calculated to estimate the posterior mean and 95% credible intervals. Posterior mode values were calculated using a kernel density estimate from the default *density()* function in R.

##### 2.3 Results

Theoretical links between Gaussian random walks, Gaussian processes, splines, and piecewise polynomials, as well as the fact that all models use the same underlying renewal model for the epidemic process, suggest that the methods should perform similarly in practice. However, we find this not to be the case.

Table S1 presents the coverage of $R_t$ and reported cases, as well as the CRPS for reported cases for each method on each of the three simulations considered in the main text. Figure S1 presents the corresponding estimates. While the methods produce mostly well-calibrated 95% uncertainty intervals for observed case data, coverage of $R_t$ estimates varies significantly, as does the CRPS for observed case incidence.

Table S1: Coverage of 95% credible intervals (confidence intervals for rtestim) for $R_t$ and observed case incidence, and CRPS for observed case incidence, for each method on each simulation.

| Method | Rt coverage | Ct coverage | CRPS |
|---|---|---|---|
| Random walk simulation | | | |
| EpiFilter (smoothing) | 93.4% | 97.8% | 4.69 |
| EpiNow2 (default) | 45.1% | 98.9% | 8.98 |
| EpiNow2 (inv-gamma) | 69.2% | 97.8% | 8.05 |
| EpiLPS (MAP) | 75.8% | 100.0% | 9.84 |
| EpiLPS (MALA) | 91.2% | 97.8% | 6.41 |
| rtestim | 100.0% | 100.0% | 10.15 |
| Sinusoidal simulation | | | |
| EpiFilter (smoothing) | 94.5% | 100.0% | 3.79 |
| EpiNow2 (default) | 73.6% | 100.0% | 14.24 |
| EpiNow2 (inv-gamma) | 76.9% | 98.9% | 5.62 |
| EpiLPS (MAP) | 91.2% | 100.0% | 9.15 |
| EpiLPS (MALA) | 92.3% | 100.0% | 4.85 |
| rtestim | 100.0% | 100.0% | 9.26 |
| Step change simulation | | | |
| EpiFilter (smoothing) | 94.5% | 100.0% | 3.10 |
| EpiNow2 (default) | 75.8% | 97.8% | 4.95 |
| EpiNow2 (inv-gamma) | 80.2% | 97.8% | 4.65 |
| EpiLPS (MAP) | 85.7% | 100.0% | 5.71 |
| EpiLPS (MALA) | 89.0% | 98.9% | 3.82 |
| rtestim | 100.0% | 100.0% | 6.08 |

The smoothed version of EpiFilter consistently outperforms (i.e. exhibits coverage of 95% credible intervals closest to 95%, and the lowest CRPS values) the other methods on these simulated datasets. This is expected in the random walk simulation, where the dynamic model employed by EpiFilter matches the simulated data. The better performance of EpiFilter on the other simulations is likely explained by two factors: (1) EpiFilter does not account for observation noise (which is absent in the simulated data), and (2) the flat prior distribution on $\eta$ is less informative than the default prior distributions used by other methods. These are both testable hypotheses that could be explored in future work to better understand the trade-offs between smoothing, model assumptions, and robustness of performance.

EpiLPS and EpiNow2 both assume that reported case incidence are negative binomially-distributed about some smooth true infection incidence (upon which the renewal model is placed), thus explicitly modelling observation noise. Process noise is derived from either the smoothness of the splines (EpiLPS) or the Gaussian process (EpiNow2). While the overdispersion parameter is estimated in both methods, the negative binomial distribution implies a lower bound of Poisson observation noise. In the simulated data, however, all noise is assumed to arise from the

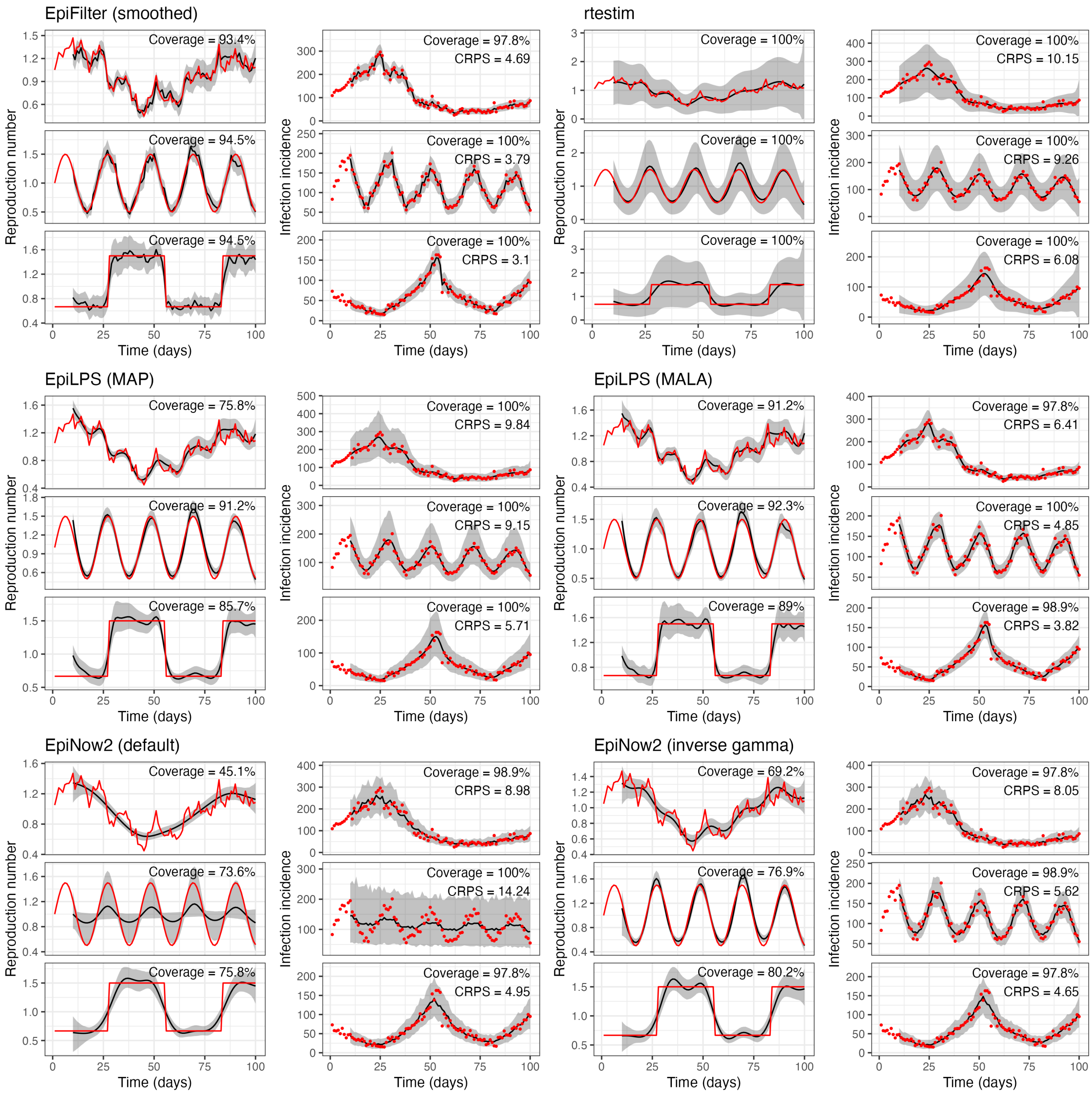

Figure S1: Estimates of $R_t$ and observed case incidence for each method on each simulation.

underlying process. By enforcing the inclusion of observation noise, EpiLPS and EpiNow2 underestimate process noise, and thus produce overly smooth estimates of $R_t$. This is likely to be less impactful on real-world datasets, where observation noise is typically a significant factor.

It is possible to use CRPS as model selection criterion instead of cross-validation. Early code to do this is provided on GitHub.

The MALA version of EpiLPS outperforms the MAP version. This is unsurprising given that, in the MALA version, uncertainty associated with $\lambda$ and $\rho$ is fully marginalised out, whereas the MAP version selects optimal point estimates of these parameters. This lends support to the argument in the main paper for the marginalisation of these parameters over selection. The MALA version also targets the true posterior distribution for $\mu(t)$, whereas the MAP version uses a Laplace approximation, which is particularly advantageous when comparing CRPS values.

Figure S1 also highlights critical oversmoothing in default EpiNow2 on the sinusoidal simulation. The posterior distribution for lengthscale $\ell$ is bi-modal, with one mode at a small value of $\ell$

Table S2: MAP and 95% credible intervals for the parameter prior and posterior distributions, where they exist. The upper portion of the table refers to smoothing parameters and the lower portion refers to estimated overdispersion. Numerical instabilities lead to large variations in statistics in some cases which are marked by a "-". As rtestim is frequentist, there are no prior assumptions on $\lambda$, and the values are chosen by cross-validation instead of being MAP estimates.

| Parameter | Prior<br>Mode (95% Cr.I) | Posterior mode and 95% Cr.I. / rtestim optimal value<br>Random walk | <br>Sinusoidal | <br>Step change |
|---|---|---|---|---|
| EpiFilter $\eta$ | U(0,1)<br>*flat* (0.025, 0.975) | 0.11 (0.08, 0.15) | 0.16 (0.13, 0.19) | 0.14 (0.11, 0.18) |
| EpiNow2 $\ell$ | LogN($\mu = 21$, $\sigma = 7$)<br>17.9 (10.5, 37.6) | 16.9 (9.64, 31.7) | 7.07 (5.33, 34.3) | 10 (6.43, 21.6) |
| EpiNow2 (inv-gam) $\ell$ | InvGam(1.5, 3.4)<br>1.38 (0.736, 31.9) | 1.35 (0.641, 7.46) | 1.99 (1.1, 4.77) | 1.77 (0.852, 6.4) |
| EpiLPS(MAP) $\lambda$ | *Hierarchical*<br>0 (0, 3.48) | 129 | 77.1 | 99.4 |
| EpiLPS(MALA) $\lambda$ | *Hierarchical*<br>0 (0, 3.48) | 103 (55.5, 380) | 76.5 (44.5, 189) | 79.9 (29.6, 468) |
| rtestim $\lambda$ | Not applicable | 1.41 | 1.41 | 1.21 |
| EpiNow2 $\phi$ | $\phi^{-1/2} \sim HalfN(0,1)$<br>- | 65.6 (42.3, 124) | 0 (7.37, 10200) | 68.5 (41.7, 253) |
| EpiNow2 (inv-gam) $\phi$ | $\phi^{-1/2} \sim HalfN(0,1)$<br>- | 86 (52.6, 199) | 0 (205, 108000) | 0 (54, 718) |
| EpiLPS(MAP) $\rho$ | $\Gamma(10^{-4}, 10^{-4})$<br>- | 25.1 | 25.2 | 25.2 |
| EpiLPS(MALA) $\rho$ | $\Gamma(10^{-4}, 10^{-4})$<br>- | 180 (84.4, 4070) | 937 (313, 17500) | 243 (105, 14200) |

(where the model follows the data) and another mode at a larger value of $\ell$, where the model estimates flat incidence with large reporting noise accounting for changes in observed cases.

##### 2.3.1 Prior distributions on smoothing parameters

While all methods listed estimate smoothing parameter(s) from the data, they handle this in different ways. Our implementation of EpiFilter, EpiNow2, and EpiLPS(MALA) marginalise this parameter out. EpiLPS(MAP) and rtestim select optimal point values of this parameter.

Of particular note is EpiNow2's prior on lengthscale $\ell$. Prior and posterior modes and credible intervals are give in Table S2. The default log-normal prior distribution often leads to over-smoothing, with an extreme example being the bimodal posterior on the sinusoidal simulation - where one mode implies that $R_t$ is (nearly) fixed, and all variation in reported cases is assigned to the observation process.

By default, we place a uniform prior distribution on EpiFilter's $\eta$ parameter. While we avoid claiming that the uniform prior distribution is uninformative, we do highlight a key advantage of this: the MAP is equal to the MLE. By the predictive decomposition of the likelihood, this is also the value of $\eta$ that optimises one-step-ahead predictions. That is, using a flat prior distribution enforces a MAP that is optimal for one-step-ahead forecasting. Furthermore, if the model is correctly specified, this implies that the MAP is optimal for any n-step-ahead forecast.

### 3 Simulated data

We test our methods on simulated data from three dynamic models.

##### Gaussian random walk

The first model is a Gaussian random walk with standard deviation $\eta\sqrt{R_{t-1}}$, matching the dynamic model assumed by EpiFilter. Simulations are initialised with default values of $R_0 = 1$, $C_{:0} = 100$, and $\eta = 0.1$. To simulate $C_{1:100}$, we iteratively sample $R_t \sim N(R_{t-1}, \eta\sqrt{R_{t-1}})$ and $C_t \sim \text{Poisson}(R_t\Lambda_t)$.

To ensure that simulations are realistic, we reject and re-simulate any simulated epidemic where daily cases exceed 5000 or fall below 5. We also truncate $R_t$ on $[0.1, 5.0]$ to ensure that results are not biased by unrealistic values of $R_t$.

##### Sinusoidal model

The second model is a sinusoidal model, which assumes that $R_t$ follows a deterministic sinusoidal function of time. Default values are a period of $\omega = 21$ days, an amplitude of $A = 0.5$, an initial value of $R_0 = 1$, and initial cases $C_0 = 100$. On day $t$, $R_t$ is assumed to be:

$$R_t^{sin} = R_0 + A \sin\left(\frac{2\pi}{\omega}t\right) \tag{S25}$$

and $C_t$ is then sampled from the Poisson renewal model with rate $R_t^{sin}\Lambda_t$.

##### Step-change model

The third model is a step-change model, which assumes that $R_t$ alternates between two values $R_a$ and $R_b$ every $T_{step}$ days. Default values are $R_a = 0.67$, $R_b = 1.5$, $T_{step} = 28$ days, and initial cases $C_0 = 100$. Depending on $t$, $C_t$ is sampled from the Poisson renewal model with rate $R_a\Lambda_t$ or $R_b\Lambda_t$.

##### Additional details

To ensure that simulations are realistic, we assume that cases prior to $t = 0$ are all equal to $C_0$. This ensures the calculation of $\Lambda_t$ is well-defined (i.e. there are past infections covering the entire generation time distribution). These assumed prior cases are not included in the simulated data, and are only used to calculate $\Lambda_t$.

While we do not consider this in the main paper, we do consider observation noise in the supplementary material. This is simulated by assuming some under-reporting rate $p$ (default $p = 0.5$), initialising the model with $C_0 = 100/p$, generating interim cases $\tilde{C}_{1:T}$ as above, and sampling $C_t \sim \text{Binomial}(\tilde{C}_t, p)$. Additional reporting noise is included by sampling from a beta-binomial distribution with $\alpha = Np$ and $\beta = N(1-p)$, where $N$ controls the amount of over-dispersion in reporting (default $N = 100$).

### 4 Serial intervals

#### 4.1 Serial interval uncertainty

We have thus far focused on uncertainty in the smoothing parameter while assuming that the serial interval is known. Here we demonstrate how these methods can be extended to also account for uncertainty in the serial interval.

Letting $\theta$ denote an arbitrary smoothing parameter and $\phi$ denote the serial interval, we target the following posterior distribution for $R_t$:

$$P(R_t|C_{1:t}) = \int P(R_t|C_{1:t}, \phi)P(\phi)\ d\phi \tag{S26}$$

Until now, we have been implicitly targeting $P(R_t|C_{1:t}, \phi)$, even if we haven't been explicitly writing $\phi$ in the conditional.

We can write this as an expectation which can be approximated by Monte Carlo methods:

$$\begin{aligned} P(R_t|C_{1:t}) &= E_\phi \left[ \int P(R_t|C_{1:t}, \phi, \theta)P(\theta|C_{1:t}, \phi)\ d\theta \right] \\ &\approx \frac{1}{N}\sum_{i=1}^{N} \left[ \int P(R_t|C_{1:t}, \phi_i, \theta)P(\theta|C_{1:t}, \phi_i)\ d\theta \right], \quad \phi_i \sim P(\phi) \end{aligned} \tag{S27}$$

A similar averaging approach is used to develop the marginal posterior distribution for predictive cases.

We demonstrate these methods by assuming that $\phi$ represents a random Gamma distribution with shape and scale parameters:

$$\alpha = 2.3669\xi, \theta = 2.7463\xi, \quad \text{where } \xi^2 \sim N(1, 0.167)$$

$E[\phi]$ is a Gamma distribution with mean 6.5 days and standard deviation 4.2 days, with 99% of the mass on a mean between 3.25 and 13 days (0.5 and 1.5 times the expected mean).

Figure S3 demonstrates the effect of applying equation S27 to the random walk simulation, using 50 draws from the random Gamma distribution described above for $\phi$. As $E[\phi]$ matches the true serial interval distribution pointwise, the error averages out, and we obtain (to Monte Carlo error), matching results whether we include uncertainty about $\phi$ or not.

The apparent irrelevance of the serial interval in this case is expected, but will not always be the case. This also relies upon $P(\phi)$ being centred on the true $\phi$. If this is not the case, then we have serial interval misspecification, which we explore in the next section.

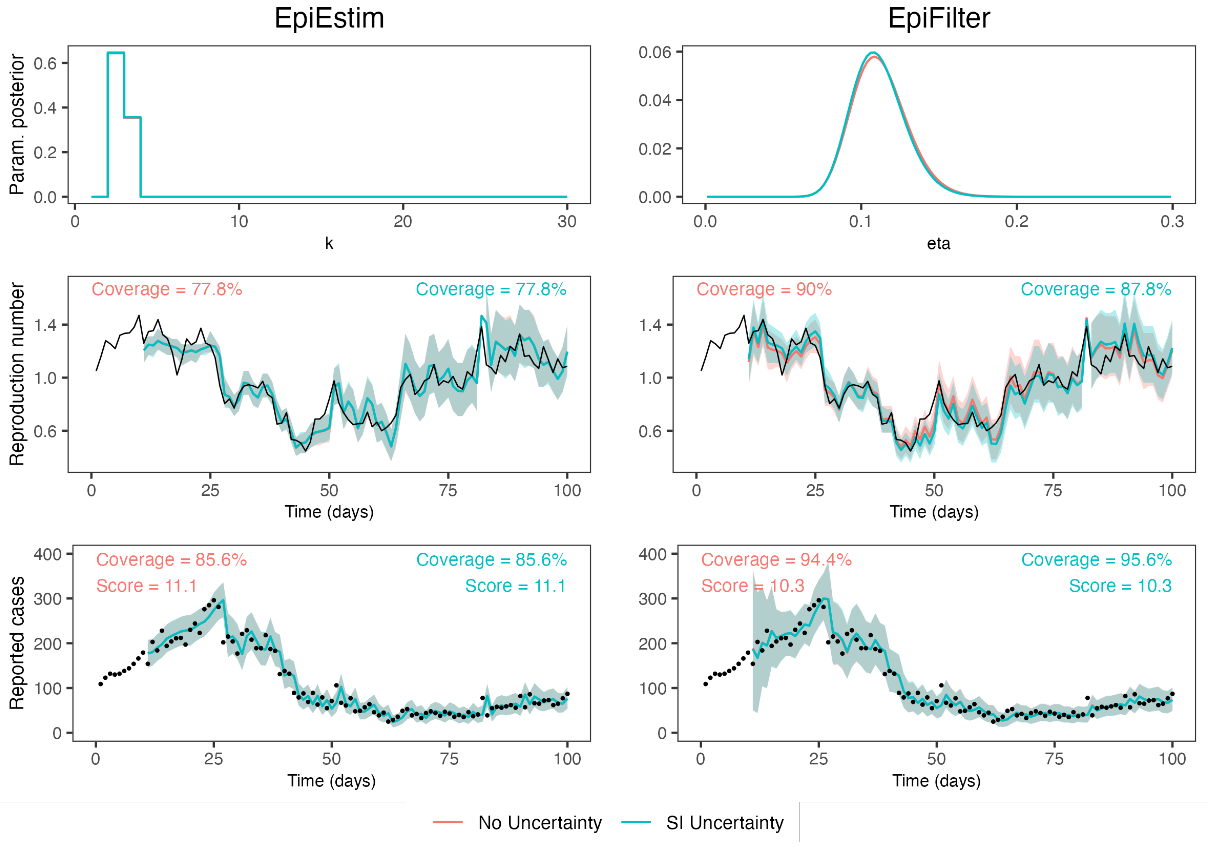

Figure S2: Results from fitting the marginalised models while also accounting for uncertainty about the serial interval. Symmetry about the expected serial interval distribution produces very similar results whether we include uncertainty about the serial interval or not.

#### 4.2 Serial interval misspecification

Serial interval misspecification is known to bias estimates of $R_t$. Even in scenarios where high quality data are available to estimate this distribution, unexpected biases can occur. During the COVID-19 pandemic, non-pharmaceutical interventions were found to shorten the generation time distribution of SARS-CoV-2 [2], and different SARS-CoV-2 variants also appeared to feature different generation times [5]. Other work has attempted to find solutions to this problem [7], we simply demonstrate the effects here.

We fit the marginalised models to the standard random walk simulation, assuming different serial interval distributions. The standard (correct) interval with mean 6.5 days and standard deviation 4.2 days is compared to results using a shorter interval with mean and standard deviation 4.2 days (an exponential distribution), and a longer interval with mean 9.75 days (1.5x the standard mean) and standard deviation 4.2 days. The interval is modified only during model fitting, the simulation itself remains unchanged.

Figure S3 and Table S3 highlight that, while the posterior distribution of the smoothing parameter depends slightly on the serial interval, and $R_t$ estimates depend strongly on the serial interval, the posterior predictive distributions are largely independent of this choice. This is an expected result, as $R_t$ and the serial interval are not jointly identifiable from reported case data alone. Diagnostics based on the predictive distribution of observed cases are unable to highlight serial interval misspecification.

Table S3: Coverage of 95% credible intervals and CRPS values for the marginalised models fit to the random walk simulation with different serial intervals.

| | Rt coverage | | Predictive coverage | | CRPS | |
|---|---|---|---|---|---|---|
| Serial interval | EpiEstim | EpiFilter | EpiEstim | EpiFilter | EpiEstim | EpiFilter |
| Standard (correct) | 77.8% | 90.0% | 85.6% | 94.4% | 11.1 | 10.3 |
| Exponential (shorter mean) | 74.4% | 86.7% | 86.7% | 94.4% | 10.8 | 10.8 |
| Longer mean | 64.4% | 72.2% | 85.6% | 96.7% | 10.8 | 10.3 |

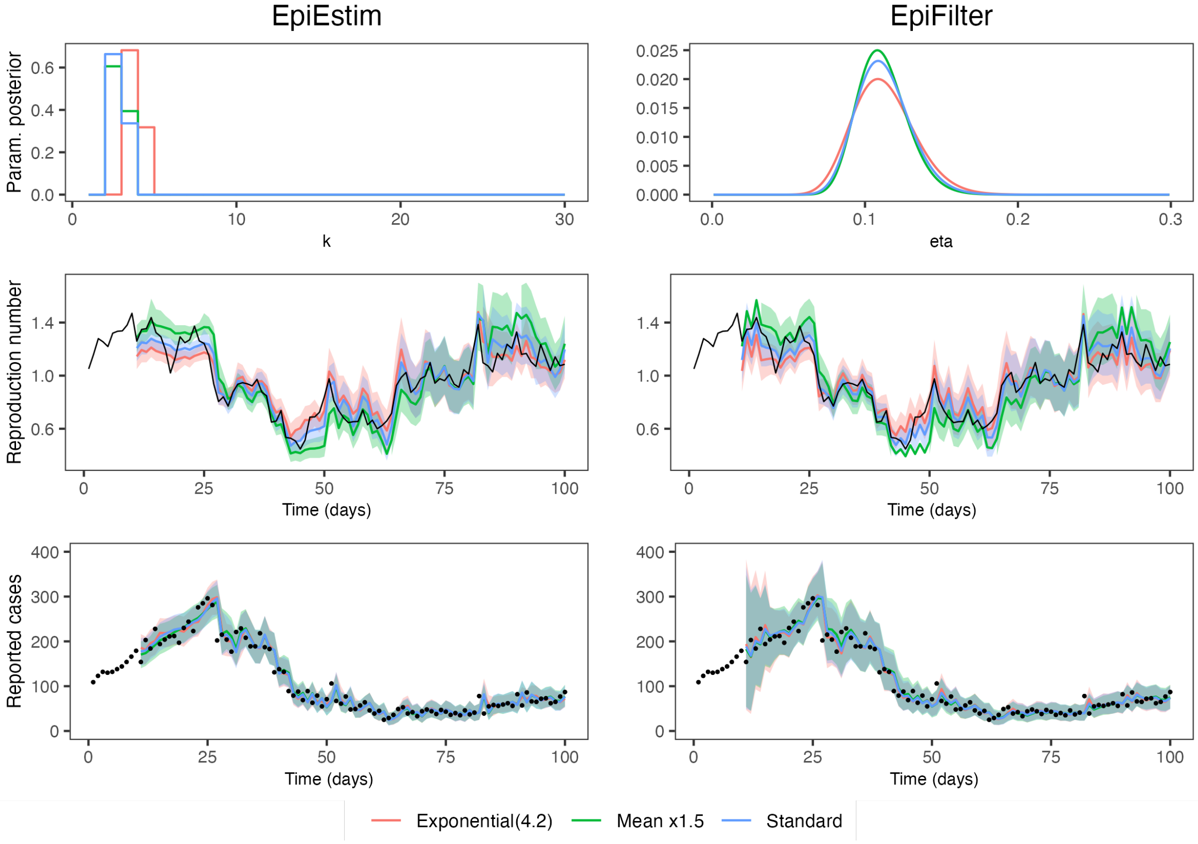

Figure S3: Results from fitting the marginalised models to the random walk simulation with different serial intervals. The standard serial interval has a mean of 6.5 days and standard deviation of 4.2 days. The exponential serial interval has a mean of 4.2 days and standard deviation of 4.2 days. The longer serial interval has a mean of 9.75 days and standard deviation of 4.2 days.

### 5 Additional simulated results

#### 5.1 Varying sample sizes

Common intuition suggests that the quality of inferences should improve as the sample size (i.e. the number of daily cases) increases. This is true for a correctly-specified model, but model misspecification, including in the choice of smoothing parameter, can lead to inferences that deteriorate with sample size (that is, the model becomes more confidently incorrect).

To demonstrate, we initialise the sinusoidal simulation with $C_0 = 25, 50, \ldots, 1600, 3200$ cases and fit the four models. Figure S4 reports the coverage of the 95% credible intervals for both $R_t$ and predictive cases, showing that while EpiFilter is largely robust to sample size, EpiEstim's coverage worsens as sample size increases in both the default and marginalised models. At a sufficiently large number of daily infections, EpiEstim's coverage increases again, as the posterior distribution of $k$ becomes more concentrated around $k = 1$, and the model returns largely unsmoothed estimates of $R_t$. This causes the CRPS score to worsen dramatically, as credible intervals become extremely wide, and the model becomes extremely under-confident in its estimates. This highlights that problems caused by incorrect smoothing cannot be solved by increasing the number of daily cases, making marginalisation a crucial tool for both small and large epidemics.

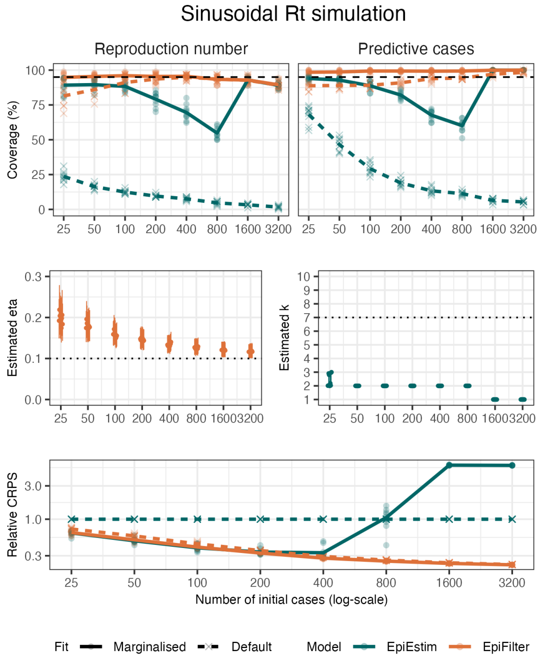

Figure S4: Coverage of the 95% credible intervals for $R_t$ and predictive cases (first row), posterior mean and 95% credible intervals for estimated parameters (second row), and the relative CRPS score (compared to default EpiEstim) for the four models fit to data from the sinusoidal simulations with varying initial number of cases $C_0$. A total of 10 simulations at each value of $C_0$ were performed and the results averaged to eliminate some stochasticity (solid lines). Results from individual simulations are reported in lighter points.

#### 5.2 Varying epidemic dynamics

We now test the models over a range of epidemic dynamics. The simulated rate of change of $R_t$ is increased by (a) increasing the standard deviation of the random walk (by increasing $\eta$), (b) decreasing the period of the sinusoidal curve, and (c) increasing the number of evenly-spaced step-changes. The rate of change of $R_t$ is decreased by doing the opposite. We fit the four models to 10 realisations of each dynamic model at each parameter value and report the average coverage, parameter estimates, and relative CRPS scores in figure S5.

As the rate at which $R_t$ varies increases, the coverage of models with default smoothing parameters decreases, as they increasingly oversmooth. Applying our marginalisation technique improves coverage, with marginalised EpiFilter being the only inference approach that achieves (approximately) correct coverage under all considered epidemic dynamics. The CRPS score is almost exclusively better for our marginalised models, except when epidemic dynamics result in the posterior distribution for EpiEstim's $k$ having a large amount of mass at $k = 1$.

Default parameter values are shown in dotted lines in the second row of figure S5, highlighting that $\eta = 0.1$ is an acceptable point estimate for EpiFilter when $R_t$ follows a sinusoidal curve with period greater than 30 days, or that $k = 7$ is an acceptable point estimate for EpiEstim when $R_t$ follows a random walk with $\eta = 0.05$, for example. Finally, the marginalisation of EpiFilter, when fit to the random walk simulation, is consistently able to recover the true value of $\eta$.

##### Practical implications

We also consider the proportion of time that each model is correctly and incorrectly confident in the sign of $R_t - 1$ (figure S6). A model is confident in epidemic growth if the 95% credible interval for $R_t$ is entirely above 1. We say it is correctly confident if the true value of $R_t$ is also greater than 1, and incorrectly confident otherwise. A similar argument applies for confidence in epidemic decline.

These results demonstrate that, while marginalisation generally increases the proportion of time in which models are correctly confident in the sign of $R_t - 1$, the crucial advantage is in reducing the proportion of time that the model is incorrectly confident in the sign of $R_t - 1$, which can be as high as 40% for default EpiEstim (in the sinusoidal $R_t$ simulation with a rapidly changing $R_t$). This has important implications for public health decision-making and communication, which often uses whether $R_t > 1$ or not to indicate the state of the epidemic and to motivate interventions.

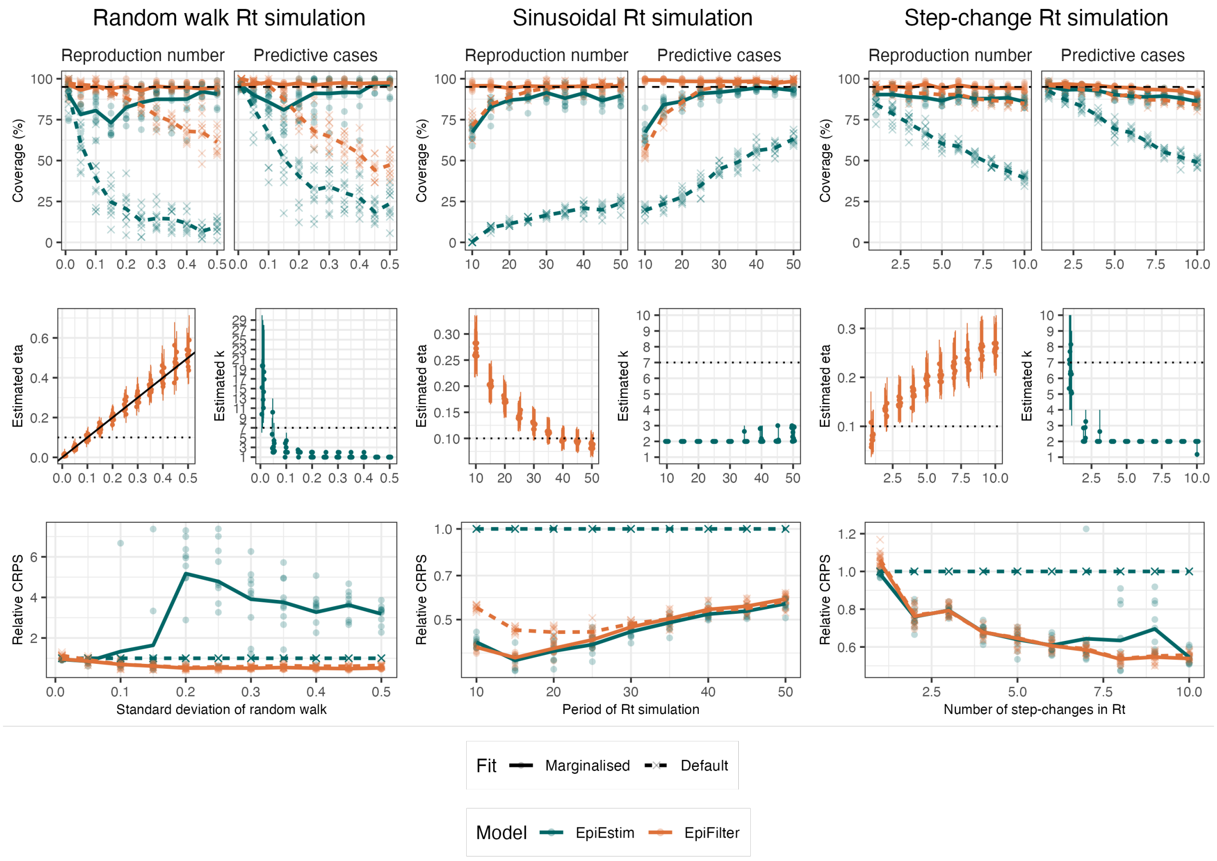

Figure S5: Coverage of 95% credible intervals for $R_t$ and predictive cases (first row), parameter estimates (second row), and relative CRPS compared to default EpiEstim (third row) for the four models fit to data from the three dynamic simulations with varying parameters. A total of 10 simulations at each parameter value were performed and the results averaged to eliminate some stochasticity (solid lines). Results from individual simulations are reported in lighter points. Default parameter values are shown in dotted lines, and the true value of the parameter (when EpiFilter is fit to the random walk simulation) is shown in a solid line.

#### 5.3 Observation-based noise

Introducing observation-based noise to the simulations in figure 2 results in posterior distributions for smoothing parameters that imply less smooth estimates (higher estimated $\eta$, lower estimated $k$), as the additional noise in the data is incorrectly assumed to arise from the epidemic process. We reproduce figure 2 with simulated observation noise in figure S7. While marginalisation in this case still improves coverage of predictive cases (particularly in EpiFilter, where coverage of predictive cases remains approximately correct in all three simulations), the same guarantee no-longer applies to $R_t$ estimates.

Comparing figure 2 to figure S7 also highlights that, while coverage of $R_t$ is generally worse in the presence of observation-based noise, the decrease is much more marked in EpiFilter than in EpiEstim. In the sinusoidal and step-change examples, when observation noise is included, marginalised EpiEstim's coverage of $R_t$ becomes comparable to that of marginalised EpiFilter's.

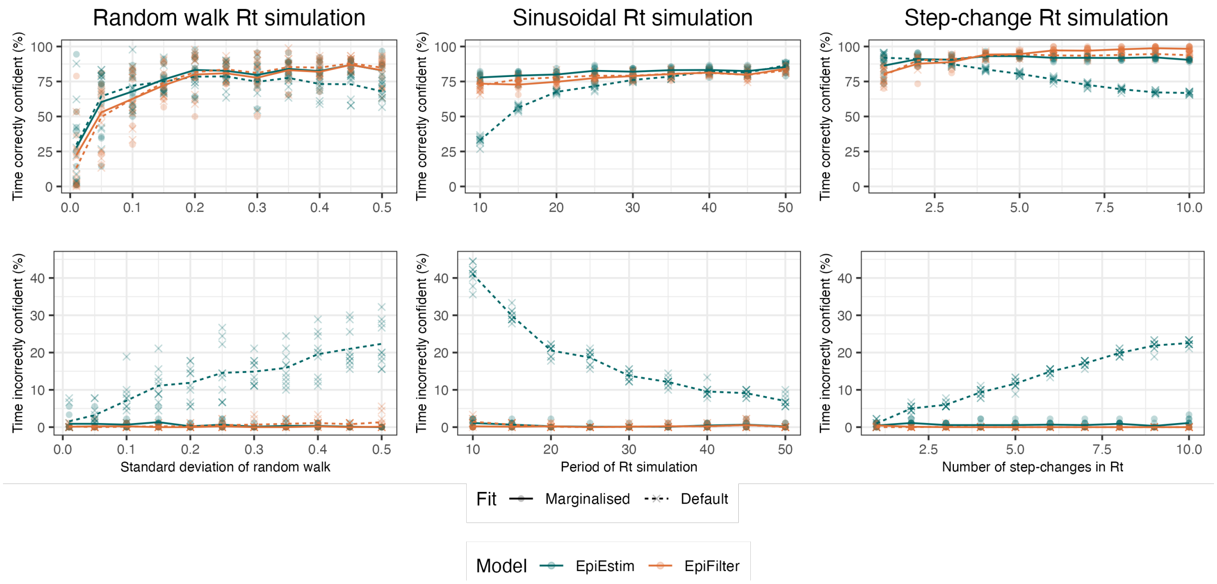

Figure S6: The proportion of time each model is correctly confident in the sign of $R_t$ (first row) and proportion of time each model is incorrectly confident in the sign of $R_t$ (second row) as a function of epidemic dynamics. A total of 10 simulations at each parameter value were performed and the results averaged to eliminate some stochasticity (solid lines). Results from individual simulations are reported in lighter points.

#### 5.4 Alternative epidemic simulations

Thus far we have tested the models on data simulated using a Poisson renewal model. In this section we test the models on data generated using a simple stochastic susceptible-infectious-recovered (SIR) model.

Simulations are initialised with $S_0 = 9950$ susceptible individuals, $I_0 = 50$ infectious individuals, and 0 recovered individuals. On each time-step $t$, a Poisson-distributed number of new infections is generated with rate $\beta I_{t-1} S_{t-1}/N$ and a Poisson-distributed number of recoveries is generated simultaneously with rate $\gamma I_{t-1}$. The theoretical instantaneous reproduction number is $R_t = \frac{\beta}{\gamma}\frac{S_{t-1}}{N}$.

We use $\beta = 1/4$ and $\gamma = 1/6.5$, implying a basic reproduction number of $R_0 = 1.625$ and a generation time distribution that follows a geometric distribution with mean 6.5 days. We use this generation time distribution when fitting our models (misspecification of the serial interval, and thus of the generation time distribution, is considered in supplementary section 4).

We fit each model twice: first on a realisation from this SIR model directly, and secondly on the same realisation with binomial noise (simulated by sampling $\tilde{C}_t \sim \text{Binomial}(2 * C_t, 0.5)$).

Results are presented in figure S8, which shows that marginalisation once again improves model results on simulations both with and without observation noise. This is unsurprising as the simple SIR model is a special case of the renewal model with an exponential generation time distribution (geometric in the discretised case presented here). One notable difference is that default EpiFilter *undersmooths* $R_t$, compared to the oversmoothing observed in other simulations. This is a result of more gradual changes in $R_t$ (the reproduction number in this SIR model changes only as a result of accumulated immunity).

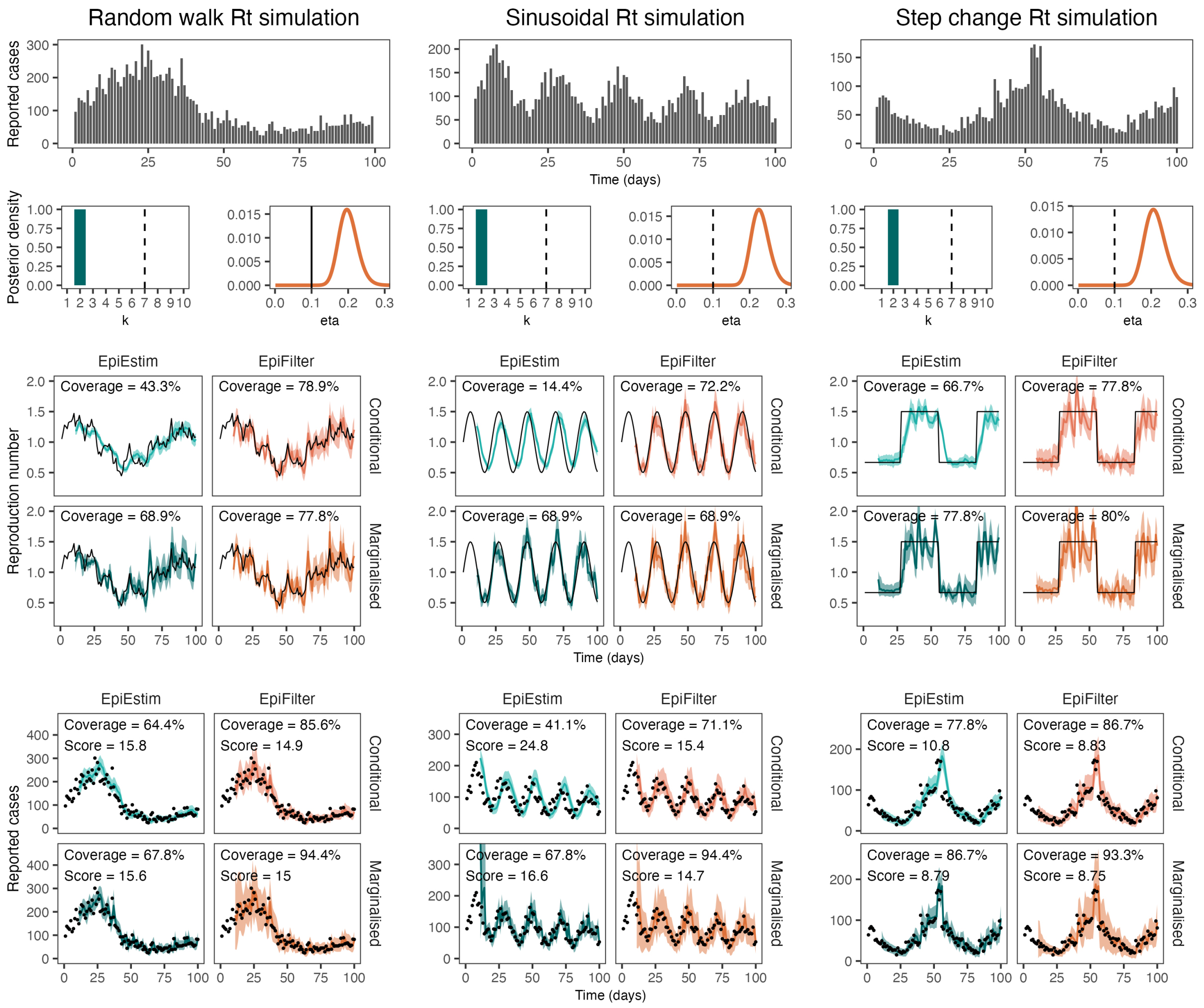

Figure S7: Simulated case data (first row), posterior distributions for smoothing parameters at $t = 100$ (second row), estimates of $R_t$ (third and fourth row), and estimates of predictive cases (fifth and sixth row) for three realisations of simulated epidemics, **with additional simulated observation noise**. The first column shows results for the Gaussian random walk simulation with $\eta = 0.1$, a dynamic model that precisely matches default EpiFilter. The second column shows results for the sinusoidal simulation, and the third column shows results for the step-change simulation. Vertical dotted lines in the parameter posterior distributions indicate the default parameter values, while the vertical solid line indicates both the default parameter value and the true value of the parameter (in EpiFilter when fit to the random walk simulation). Black lines (in $R_t$ estimates) and black dots (in predictive $C_t$ estimates) show the true values of $R_t$ and $C_t$ respectively. Predictive coverage of the 95% credible intervals (closer to 95% is better) and the CRPS scores (lower is better) are shown within each figure.

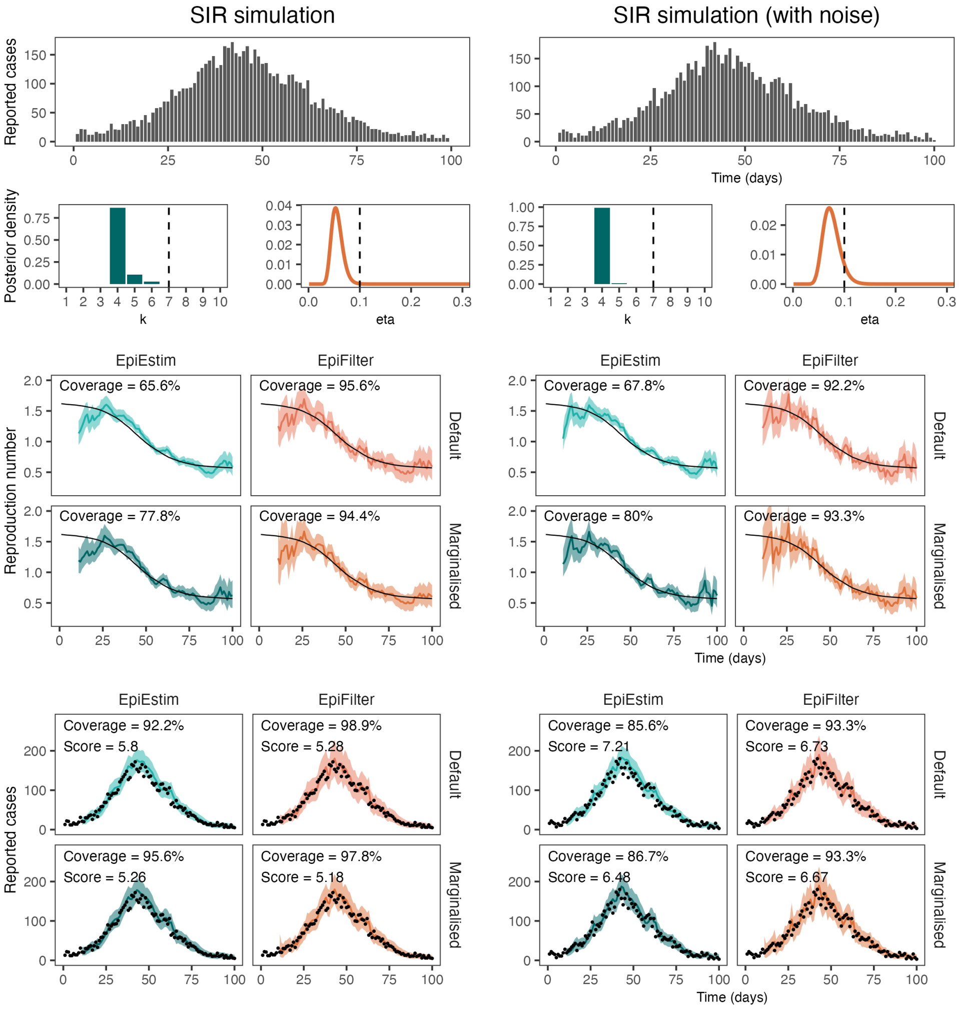

Figure S8: Simulated case data (first row), posterior distributions for smoothing parameters at $t = 100$ (second row), estimates of $R_t$ (third and fourth row), and estimates of predictive cases (fifth and sixth row) for two realisations of simulated epidemics from an SIR model. The first column shows results for the SIR simulation without observation noise, and the second column shows results for the SIR simulation with observation noise. Vertical dotted lines in the parameter posterior distributions indicate the default parameter values. Black lines (in $R_t$ estimates) and black dots (in predictive $C_t$ estimates) indicate the true values of $R_t$ and $C_t$ respectively. Predictive coverage of the 95% credible intervals (closer to 95% is better) and the CRPS scores (lower is better) are shown within each figure.

### 6 Additional real-world examples

#### 6.1 New Zealand data

Neither EpiEstim nor EpiFilter allow for observation noise in their models, so all noise in the data is assumed to arise from the epidemic process. In practice, reporting noise can be a significant factor. We attempt to side-step this problem when fitting to New Zealand data by imposing a 5-day moving average on the raw case data. We test the impact of this on our conclusions here by re-fitting the model on the raw data, and on data smoothed using a 10-day moving average.

Figure S9 shows that parameter estimates are highly dependent on any pre-smoothing performed on the data, with smoother estimates being produced on data that underwent more pre-smoothing. Despite this, marginalisation still consistently improves coverage of observed data and CRPS, and the models obtain good coverage of observables in general.

If observation noise is considered substantial, we encourage the reader to use a model that explicitly accounts for this. Examples in the literature that jointly handle observation noise and smoothing parameters are EpiNow2 and EpiLPS(MALA), both of which are outlined in supplementary section 2.

#### 6.2 Alternative datasets

Using the same data as [8], we present additional examples of the fitting of EpiEstim and EpiFilter to real-world data: the 1918 influenza outbreak in Baltimore and 2003 SARS outbreak in Hong Kong. The models are also fit to data that has been smoothed using a 7-day moving-average to eliminate some observation noise, which our models are unable to account for. Results are presented in figure S10 which shows that marginalisation almost universally improves both the coverage of predictive cases and the CRPS, with the sole exception of fitting EpiFilter to the smoothed 1918 influenza data.

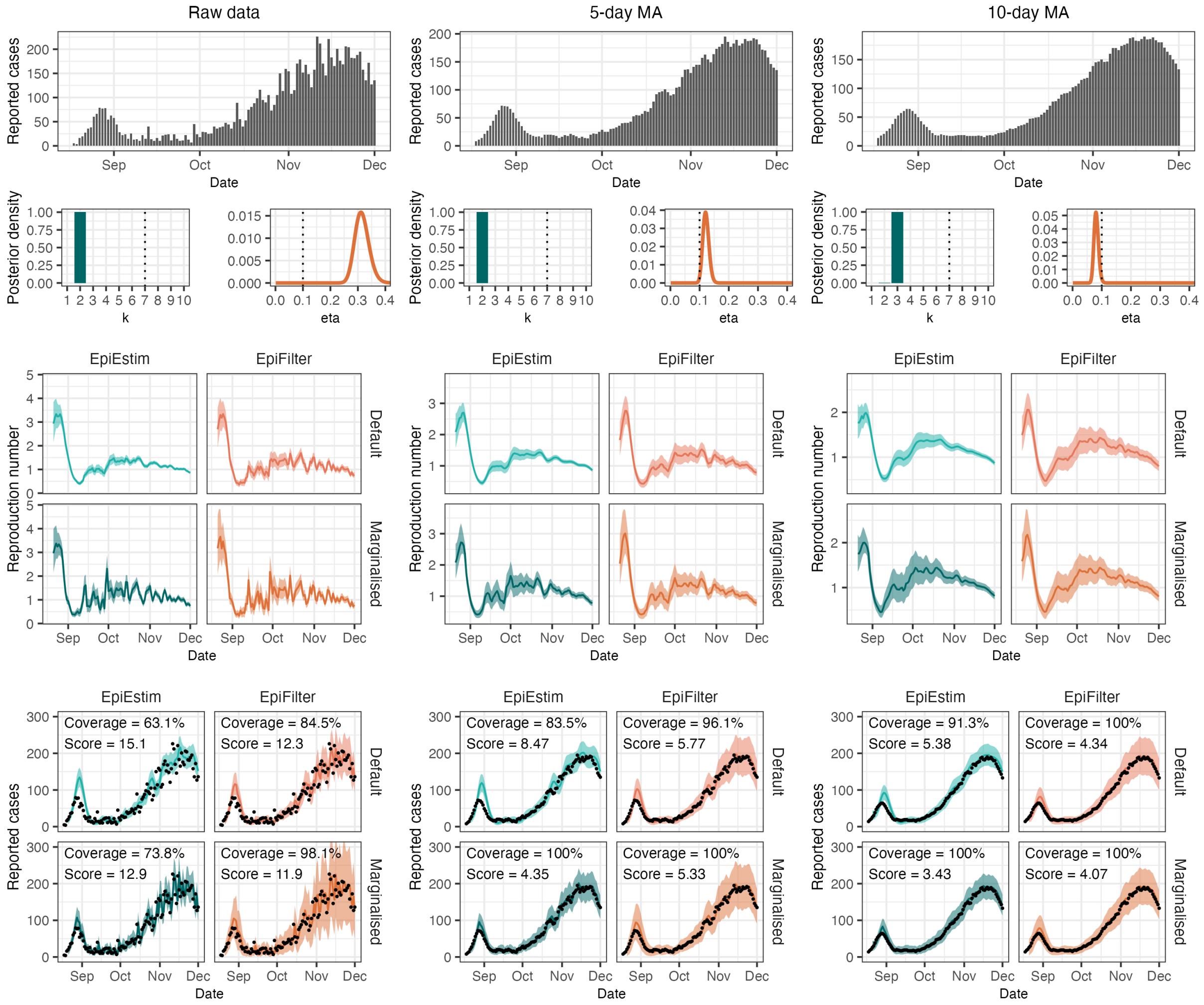

Figure S9: Results from fitting all four models to raw New Zealand data (column 1), and New Zealand data smoothed using a 5-day and 10-day moving average (columns 2 and 3 respectively). In row 2, vertical dashed lines indicate default parameter values while coloured curves show parameter posterior distributions. Coloured lines in the remaining figures show central estimates (mean of the posterior distributions) and coloured bands show 95% credible intervals. Observed case data are shown in black points.

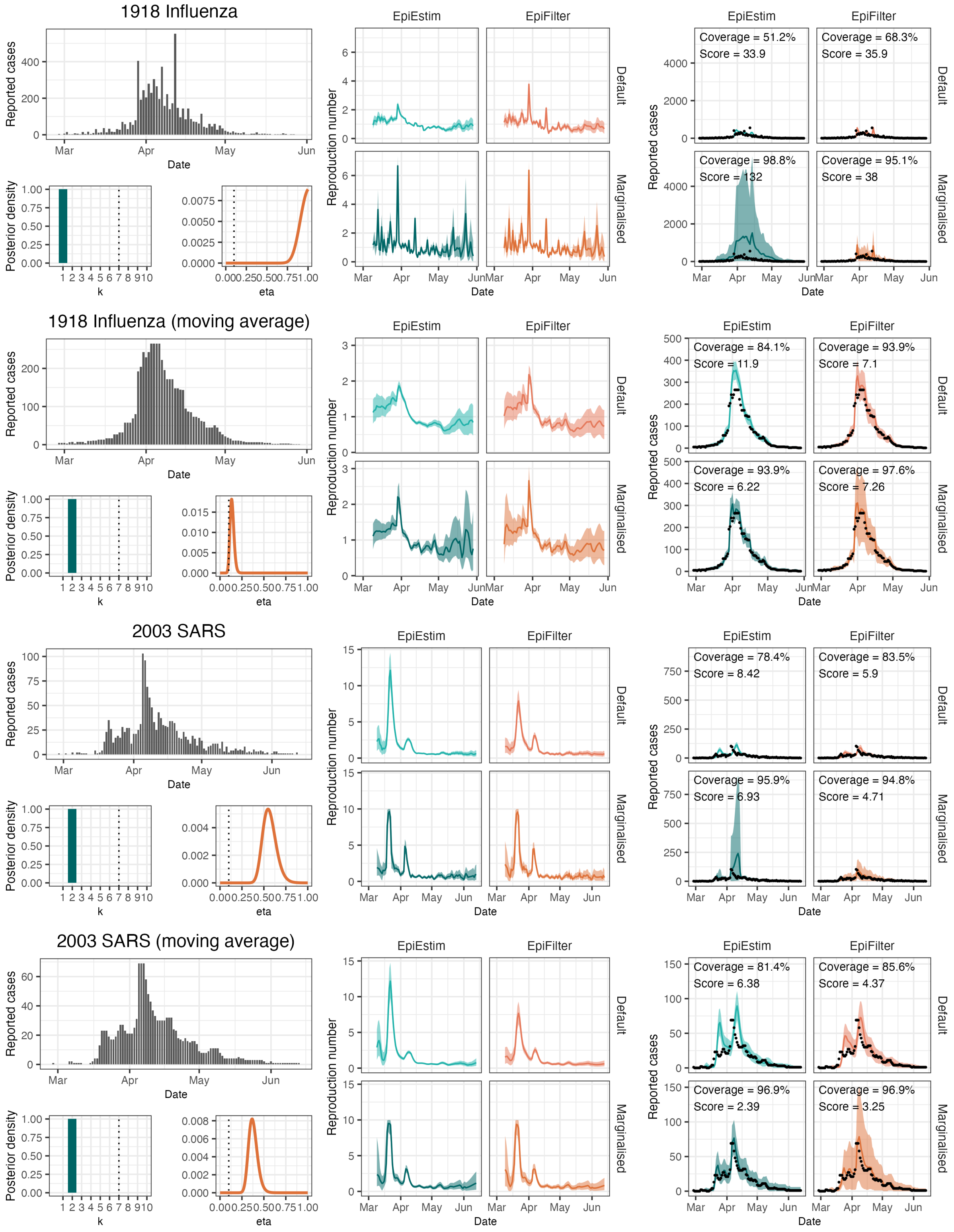

Figure S10: Results from fitting EpiEstim and EpiFilter to two real-world time-series. The models are also fit on smoothed data (to eliminate some observation noise), demonstrating that this can impact parameter estimates.

### 7 Stepwise likelihoods

The model likelihoods are calculated by summing the log-likelihood of the one-step-ahead forecasts at each time-step. By plotting these one-step-ahead likelihoods for various values of $k$ and/or $\eta$, we can identify the periods of the epidemic that are driving the observed parameter estimates (figure S11).

Most notably, we can see that, for most periods of time, smaller values of $\eta$ are preferred to larger values. However, when $R_t$ is changing rapidly (such as when $R_t$ is crossing 1 in the sinusoidal simulation), these small values incur a large "penalty" in the likelihood, resulting in more posterior mass at larger values of $\eta$. The same effect also occurs in EpiEstim, although as most mass is concentrated around small values of $k$, the effect is less pronounced.

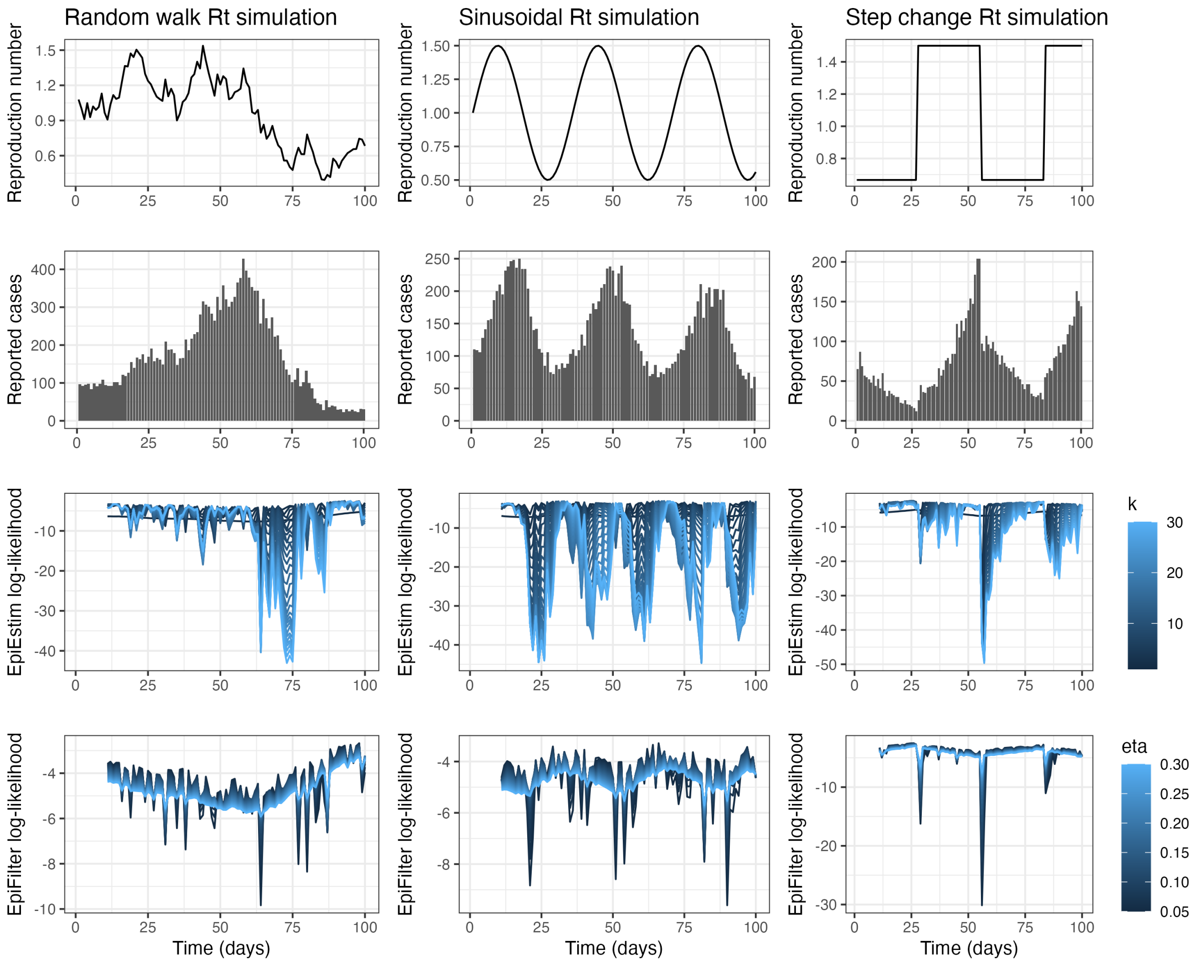

Figure S11: Simulated $R_t$ (first row), simulated reported cases (second row), stepwise log-likelihoods for EpiEstim (third row) and stepwise log-likelihoods for EpiFilter (fourth row) for the sinusoidal simulation. Darker lines show stepwise log-likelihoods associated with smaller values of $\eta$ (increased smoothing) and smaller values of $k$ (increased smoothing).

### 8 APE comparison

Parag & Donnelly [8] treat the selection of EpiEstim's $k$ as a model-selection problem and use information theoretic arguments to suggest that the optimal value of $k$ is that which minimises the accumulated prediction error (APE). The APE is defined (in our notation) as:

$$APE_k = \sum_{s<t} -\log P_{APE}(C_{s+1}|C_{1:s}, k) \tag{S28}$$

Noting the following equivalence to the predictive decomposition of the likelihood (equation S2):

$$\ell(k|C_{1:t}) = \sum_{s=1}^{t} \log P(C_s|C_{1:s-1}, k) \ \stackrel{?}{=} \ -APE_k \tag{S29}$$

The $k$ that minimises the APE should be the same $k$ that maximises the log-likelihood. The two approaches differ, however, in their derivation of $P(C_s|C_{1:s-1}, k)$.

Parag & Donnelly derive the predictive distribution for $C_s$ given $C_{1:s-1}$ by first noting that $R_{s-1}|C_{1:s-1}, k$ is Gamma-distributed with shape parameter $\alpha_{s-1,k}$ and rate parameter $\beta_{s-1,k}$, and then assuming that $C_s|R_{s-1}, \Lambda_s$ is Poisson distributed with rate $R_{s-1}\Lambda_s$. **We note that this introduces a new assumption in the model: specifically, that before we observe $C_s$, we assume $R_s$ has been fixed for the preceding $k+1$ days.** The resulting predictive distribution for $C_s$ is then a negative binomial distribution:

$$P_{APE}(C_s|C_{1:s-1}, k) \sim \text{NegativeBinomial}\left(r = \alpha_{s-1,k}, p = \frac{\beta_{s-1,k}}{\Lambda_s + \beta_{s-1,k}}\right) \tag{S30}$$

This is slightly different to our approach (equation S7), where $r = \alpha_{s-1,k-1}$ and $p = \beta_{s-1,k-1}/(\Lambda_s + \beta_{s-1,k-1})$. The $APE_k$ metric using the predictive distribution derived by Parag & Donnelly is precisely equal to $-\ell(k+1|C_{1:t})$. The additional assumption made by the APE approach results in a bias towards smaller values of $k$.

We present the APE-metric and negative-log-likelihood for values of $k$ between 1 and 30 on the New Zealand COVID-19 dataset in figure S12. We note that the APE-metric is minimised at $k = 1$, whereas the negative-log-likelihood is minimised at $k = 2$.

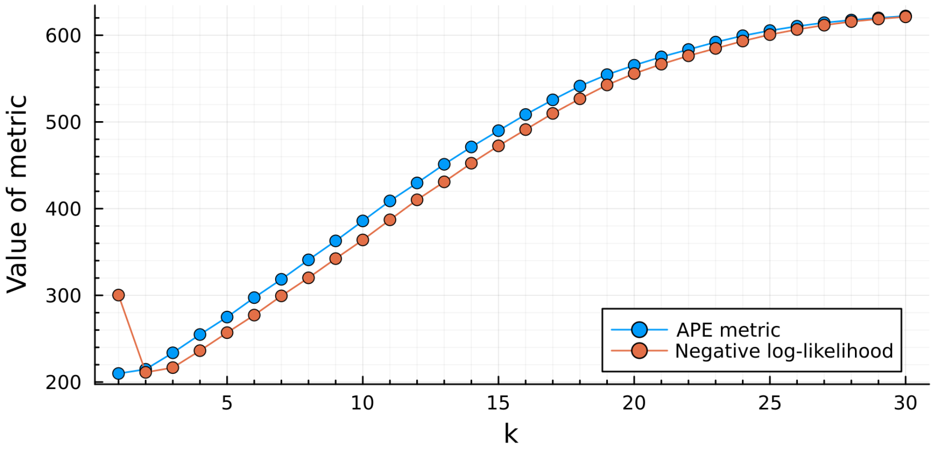

Figure S12: Comparison of the APE metric from Parag & Donnelly [8] and the negative log-likelihood of $k$ for EpiEstim on the NZ COVID-19 dataset. For $k \geq 1$, the value of $APE_k$ is exactly the same as $-\ell(k+1|C_{1:t})$. The value of $\ell(1|C_{1:t})$ is the log-likelihood of $k = 1$ (where the predictive distribution for $R_t$ is simply the prior distribution) and has no immediate analogue in the APE approach.

### 9 Testing grid-sizes

There are three instances in which we employ non-exact grid-based approximations to posterior distributions:

1. When marginalising out $k$ in EpiEstim (grid-approximation for $R_t$)
2. Both default and marginal EpiFilter (grid-approximation for $R_t$)
3. When fitting $\eta$ in EpiFilter (grid-approximation for $\eta$)

By default, these grids are chosen to be of size 1000 (supplementary material section 1.1.2). We test the sensitivity of our results to grids of size 500 (a coarser grid) and 2000 (a finer grid) and find that these grids have very little impact on our results. We further test EpiEstim with an $R_t$ grid-size of just 100, at which size there is evidence of some degeneracy in the posterior distribution for $R_t$. Full results are discussed in the captions of figures S13, S14, and S15.

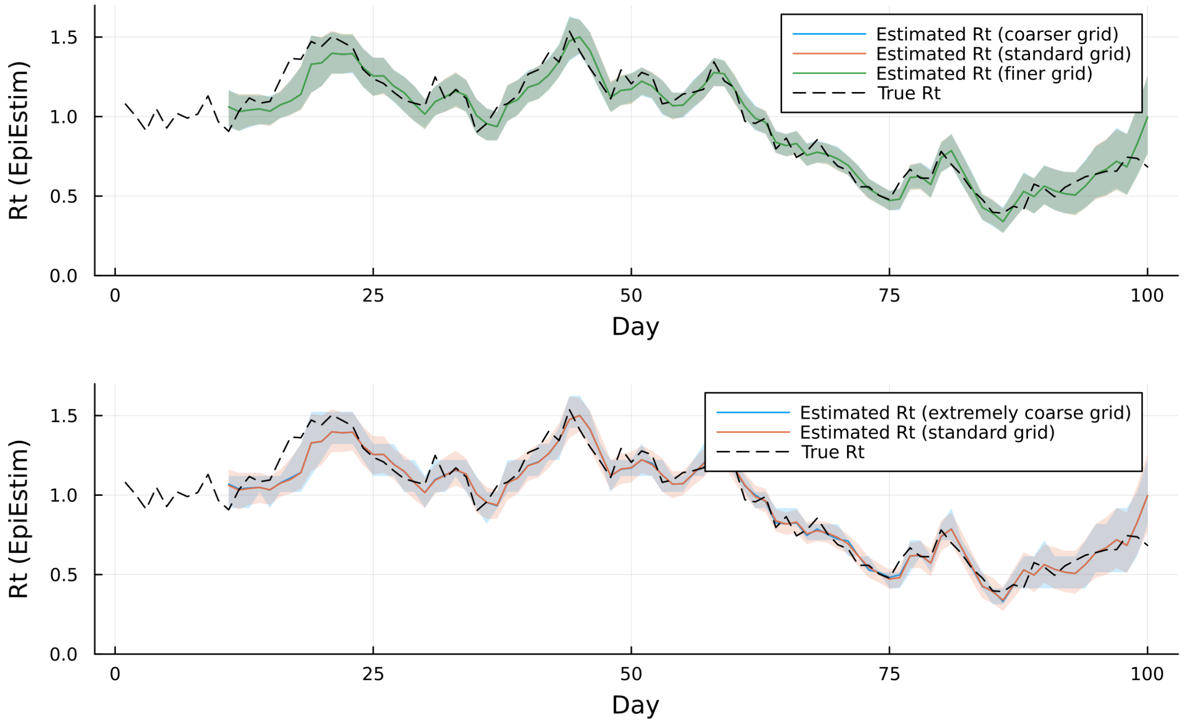

Figure S13: Results from fitting (marginalised) EpiEstim to simulated data using three different grid-sizes for $R_t$: 500 (coarser), 1000 (standard), 2000 (finer). We also test the model with a grid-size of 100 (very coarse grid). Only in the case of the very coarse grid do we see evidence of degeneracy in the posterior distribution for $R_t$, suggesting that a grid size of 500 or more is sufficient.

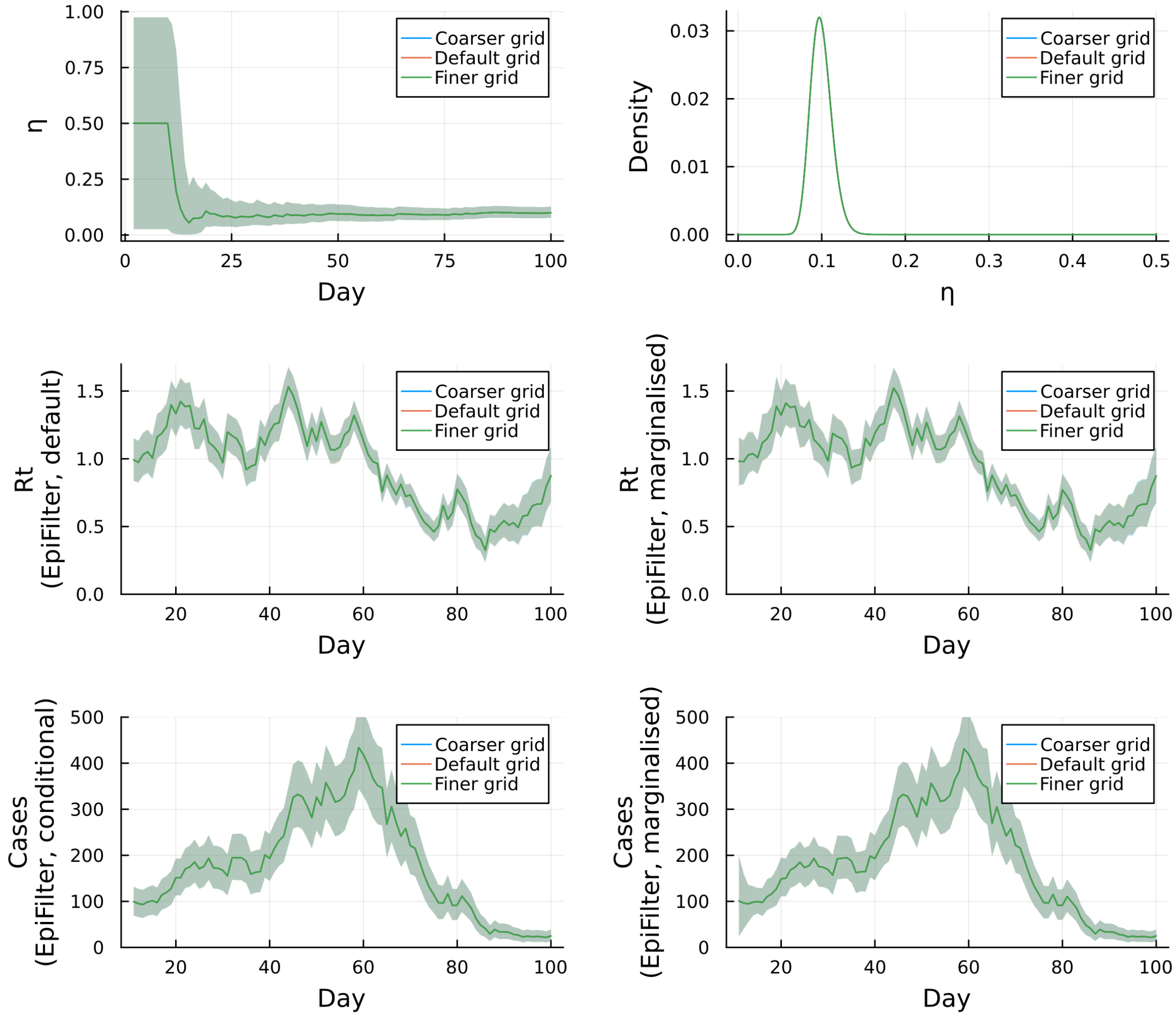

Figure S14: Results from fitting EpiFilter (default and marginalised) to simulated data using three different grid-sizes for $R_t$: 500 (coarser), 1000 (standard), 2000 (finer). We find that the choice of grid-size has very little impact on the results. *The density plot for $\eta$ is the posterior distribution of this parameter at the final time-step (i.e. conditional on all data).

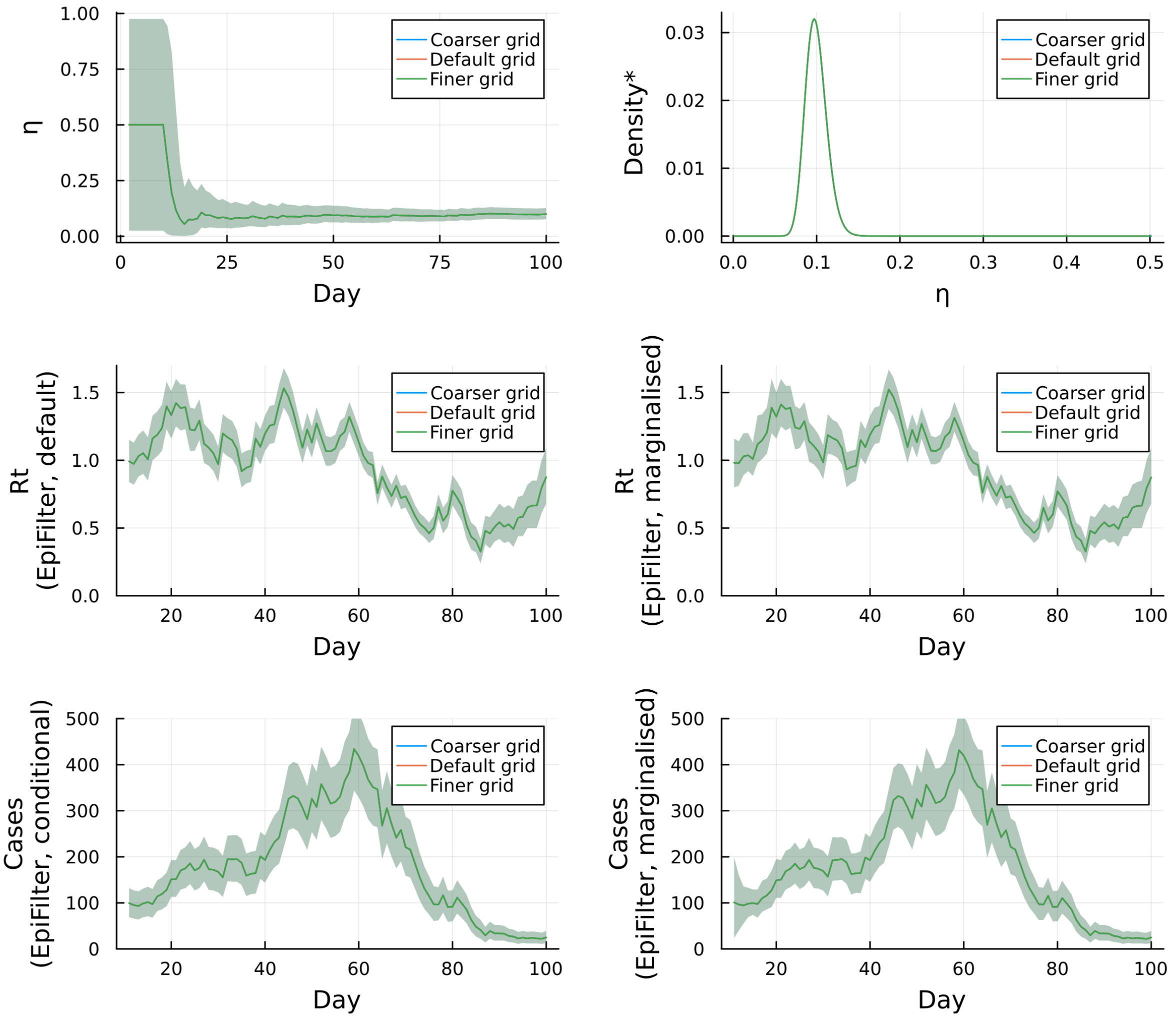

Figure S15: Results from fitting EpiFilter to simulated data using three different grid-sizes for $\eta$: 500 (coarser), 1000 (standard), 2000 (finer). We find that the choice of grid-size has very little impact on the results, such that the resulting posterior distributions are indistinguishable. *The density plot for $\eta$ is the posterior distribution of this parameter at the final time-step (i.e. conditional on all data).
